# Integrating Histopathology Foundation Models and PRAME Immunohistochemistry in Melanoma Diagnosis

**DOI:** 10.64898/2026.11.14.26368089

**Authors:** Gellrich Frank Friedrich, Kistner Alaska, Günther Claudia, Hufnagel Cosima, Hobelsberger Sarah, Laske Jörg, Beissert Stefan, Aust Daniela, Baretton Gustavo, Reidow Nick, Sergon Mildred, Kather Jakob Nikolas, Ghaffari Laleh Narmin

## Abstract

**Background:** The histopathological diagnosis of challenging melanocytic lesions remains limited by substantial interobserver variability. Although computational pathology foundation models demonstrated promising diagnostic performance, their predictions are often difficult to interpret in terms of established criteria.

**Objective:** We evaluated a diagnostic framework combining foundation-model-based whole-slide image analysis, automated prediction of morphological features, and manually assessed PRAME immunohistochemistry.

**Methods:** Using the Virchow2 foundation model and STAMP framework, we analyzed a retrospective cohort of 1,000 melanocytic lesions undergoing PRAME immunohistochemistry. Models were trained on 800 cases and evaluated on a temporally held-out test cohort of 200 cases.

**Results:** The slide-level malignancy classifier achieved a test AUROC of 0.970 (AUPRC 0.984). Separate models predicted established histopathological criteria with high discrimination, including ulceration (AUROC 0.991) and dermal mitoses (AUROC 0.966). In contrast, predicting PRAME expression from H&E images alone showed lower performance (AUROC 0.776-0.780 for PRAME scores 4 and 0). An integrated model combining the slide-level malignancy score, AI-predicted morphological features, and manual PRAME status achieved an AUROC of 0.972 (AUPRC 0.975), comparable to the standalone classifier. In multivariable analysis, PRAME score 4 versus 0 was strongly associated with malignancy (adjusted OR 35.04, 95% CI 17.39-74.62).

**Conclusions:** These findings demonstrate the feasibility of combining foundation-model-based histopathological analysis with established morphological criteria and PRAME immunohistochemistry. Although the integrated framework did not substantially improve overall discrimination, it provides a structured approach for incorporating complementary histopathological and immunohistochemical information into AI-assisted assessment of melanocytic lesions.

**Key Points:** *Why was the study undertaken?:* Deep learning models in histopathology often lack diagnostic transparency. Existing frameworks rarely incorporate established morphological criteria or manually assessed findings, such as PRAME immunohistochemistry, in the evaluation of challenging melanocytic lesions.

*What does this study add?:* We evaluated a framework combining foundation-model-based whole-slide analysis with AI-predicted morphological features and manual PRAME status. Although overall diagnostic performance was comparable to the standalone model, integrating these criteria as statistically significant predictors aligns the algorithmic output with established histopathological parameters.

*What are the implications of this study for disease understanding and/or clinical care?:* This study demonstrates the feasibility of combining computational models with standard histopathological workflows. Correlating algorithmic predictions with established criteria facilitates potential clinical applications, including automated case triaging and diagnostic decision support.

**Plain Language Summary:** Diagnosing melanoma, a type of skin cancer, under a microscope can be challenging. In difficult cases, doctors may sometimes reach different conclusions. Artificial intelligence (AI) computer programs can accurately analyze digitized tissue samples, but they often function as “black boxes,” making it difficult for doctors to understand how the AI reached its conclusion.

In this study, we tested an AI system on 1,000 skin samples. Instead of relying solely on the AI to provide a final diagnosis, we combined its analysis with specific microscopic features that doctors routinely look for, alongside the results of a special laboratory test called PRAME.

We found that the AI system was highly accurate in identifying melanoma. While combining the AI with the specific tissue features and the PRAME test did not further increase the overall accuracy, it successfully aligned the computer’s output with established medical standards.

This approach demonstrates that advanced computer models can be integrated with traditional laboratory tests. By linking AI results to familiar medical criteria, the system’s output becomes more transparent for physicians, which could help doctors evaluate difficult skin tissue samples more efficiently in the future.

## Introduction

Malignant melanoma represents one of the most aggressive and lethal forms of skin cancer ^1,2^. While histopathological examination remains the established gold standard for diagnosis and staging ^1,3,4^, the microscopic evaluation of melanocytic lesions is inherently subjective, relying on cellular atypia, architectural asymmetry, and pagetoid spread. Particularly in borderline lesions, this results in significant inter-observer variability even among experienced dermatopathologists ^5,6^, potentially leading to overdiagnosis or fatal underdiagnosis ^5^.

To address these diagnostic challenges, the digitization of histological tissue sections into whole-slide images (WSIs) has enabled computational pathology ^7,8^. Early deep learning approaches employed supervised convolutional neural networks (CNNs), requiring time-consuming manual annotations that limited scalability and generalizability across clinical datasets ^9–11^. To circumvent this constraint, weakly supervised multiple instance learning (MIL) was introduced, dividing WSIs into smaller patch instances to predict malignancy directly from slide-level diagnostic labels without manual annotations ^12–15^.

A major advance in computational pathology has been the introduction of histopathology foundation models ^16,17^. Those models are pretrained on large, diverse datasets using self-supervised learning to acquire generalizable visual representations ^18–21^. Models such as Virchow2 extract high-dimensional feature embeddings from image patches, which are aggregated by task-specific models to generate slide-level predictions ^20,22^. Coupled with downstream aggregator frameworks like STAMP (Solid Tumor Associative Modeling in Pathology), these models demonstrate high diagnostic accuracy in predicting melanoma across WSIs ^20,23^.

However, foundation models operate primarily as ‘black boxes’, providing probabilistic scores without a clinically transparent rationale ^24^. For AI to be safely adopted in routine dermatopathology, clinical interpretability is essential ^25,26^. One approach is to assess whether learned image representations can predict established histopathological criteria, such as dermal mitoses, ulceration, or junctional atypia, alongside the primary malignancy classifier ^27^. Recent studies confirmed that deep learning models reliably assess such histological structures with accuracies exceeding 92% ^28^, bridging machine learning and human evaluation to support human-in-the-loop workflows ^11,27^.

While deep learning can infer molecular alterations directly from standard H&E slides if distinct morphological correlates exist ^29^, automated digital quantification of dedicated immunohistochemical stains, such as PRAME (Preferentially Expressed Antigen in Melanoma), substantially reduces interobserver discordance compared with manual scoring ^30^. In challenging melanocytic lesions, PRAME has emerged as a sensitive and specific diagnostic tool ^31^. However, the reliability of inferring PRAME expression from routine morphology remains uncertain in borderline lesions. Integrating PRAME as an independent input therefore provides non-redundant information that complements automated image analysis and improves model robustness across cohorts ^32,33^.

The objective of this study was to evaluate a diagnostic framework combining foundation-model-based WSI analysis with manually assessed PRAME immunohistochemistry in melanocytic lesions. We first assessed the diagnostic performance of a slide-level malignancy classifier and evaluated whether foundation-model representations could predict established histopathological criteria and immunohistochemical markers. We subsequently examined the performance of an integrated model combining the slide-level malignancy score, AI-predicted morphological features, and manually assessed PRAME status.

## Material and Methods

### Patient Selection and Cohort Definition

The study design, digitization, computational workflow, and evaluation strategy are illustrated in Figure 1. This retrospective, single-center study at the Department of Dermatology, University Hospital Carl Gustav Carus, TU Dresden, identified all melanocytic lesions with PRAME immunohistochemistry selectively requested for diagnostically challenging cases between January 1, 2020, and January 21, 2025. From 1,360 consecutive cases, non-melanocytic lesions, re-excision specimens, mucosal or uveal melanomas, and metastases were excluded to restrict the dataset to primary cutaneous melanocytic neoplasms. All corresponding hematoxylin and eosin (H&E), PRAME, and routine immunohistochemical slides (SOX10, MelanA, HMB45, Ki-67), were digitized using an Aperio AT2 scanner (Leica Biosystems, Wetzlar, Germany) at 40x magnification.

**Figure 1:**
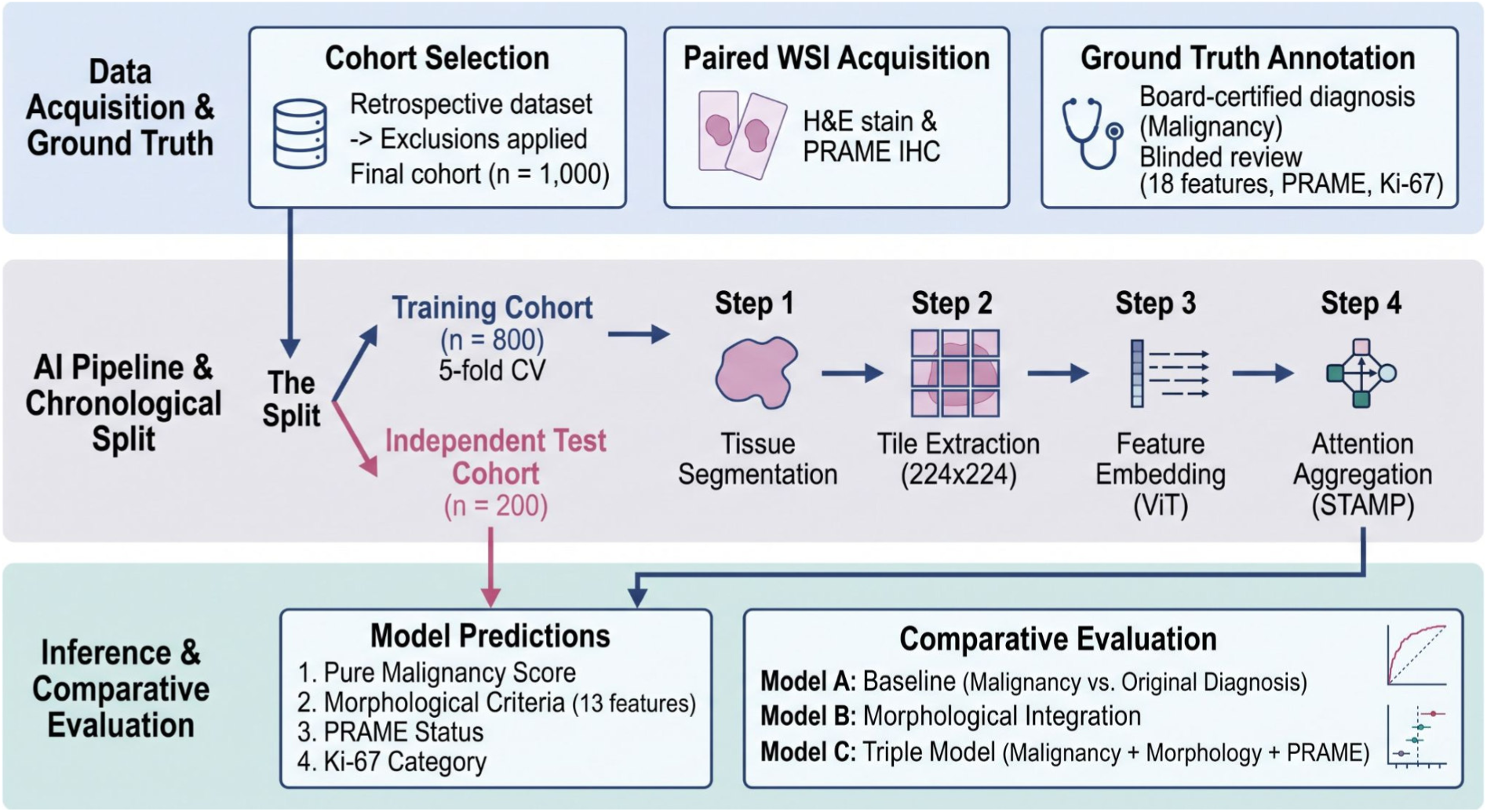
Study design, AI pipeline, and comparative evaluation framework. A retrospective cohort of 1,000 melanocytic lesions with paired H&E and PRAME IHC whole-slide images was chronologically partitioned into a training and an independent test set. The deep learning pipeline processed H&E slides via tissue segmentation, tile extraction, Vision Transformer (ViT) feature embedding, and STAMP attention aggregation. Model predictions were evaluated across three configurations: a slide-level baseline (Model A), a morphologically integrated framework (Model B), and the multimodal Triple Model (Model C). CV = cross-validation. H&E = haematoxylin and eosin. IHC = immunohistochemistry. PRAME = preferentially expressed antigen in melanoma. STAMP = Solid Tumor Associative Modeling in Pathology. ViT = vision transformer. WSI = whole-slide image.

The final cohort comprised 1,000 H&E WSIs from primary cutaneous melanocytic lesions. Multiple distinct lesions were excised in 87 patients, and multiple sections were digitized for 44 specimens. Each digitized section was treated as an independent slide-level observation. Patient age, sex, diagnostic category, PRAME status, and all 18 histological criteria were documented for all 1,000 WSIs, anatomical location in 988 cases, and Ki-67 expression in 316 stained WSIs. Due to fragmented historical reporting, conventional melanoma subtypes and chronic sun damage were not included.

In 19 WSIs exhibiting exclusively junctional features despite an original report documenting a dermal component on other section levels, the slide-level diagnostic ground truth was adjusted from invasive melanoma to melanoma in situ to align with the digitized section, leaving binary malignancy unaffected.

### Reference Standard and Study Annotation

The reference diagnosis was established from original histopathology reports issued by board-certified dermatopathologists. All malignant diagnoses (melanoma in situ and invasive melanoma) underwent mandatory institutional second-opinion review, with borderline cases resolved by interdisciplinary consensus. A single investigator, blinded to the reference diagnosis, systematically evaluated the 18 histological criteria, PRAME scores, and Ki-67 proliferation index. While all 18 criteria were utilized for baseline characterization (Table 1) and feature concordance (Supp. Tables S1, S3), a curated core set of 13 tumor-specific features was selected for machine-learning benchmarking and multivariable integration, excluding non-specific infiltrates (dense infiltrates, TILs), anatomical compartments (junctional/dermal components), and subcutis infiltration.

**Table 1:**
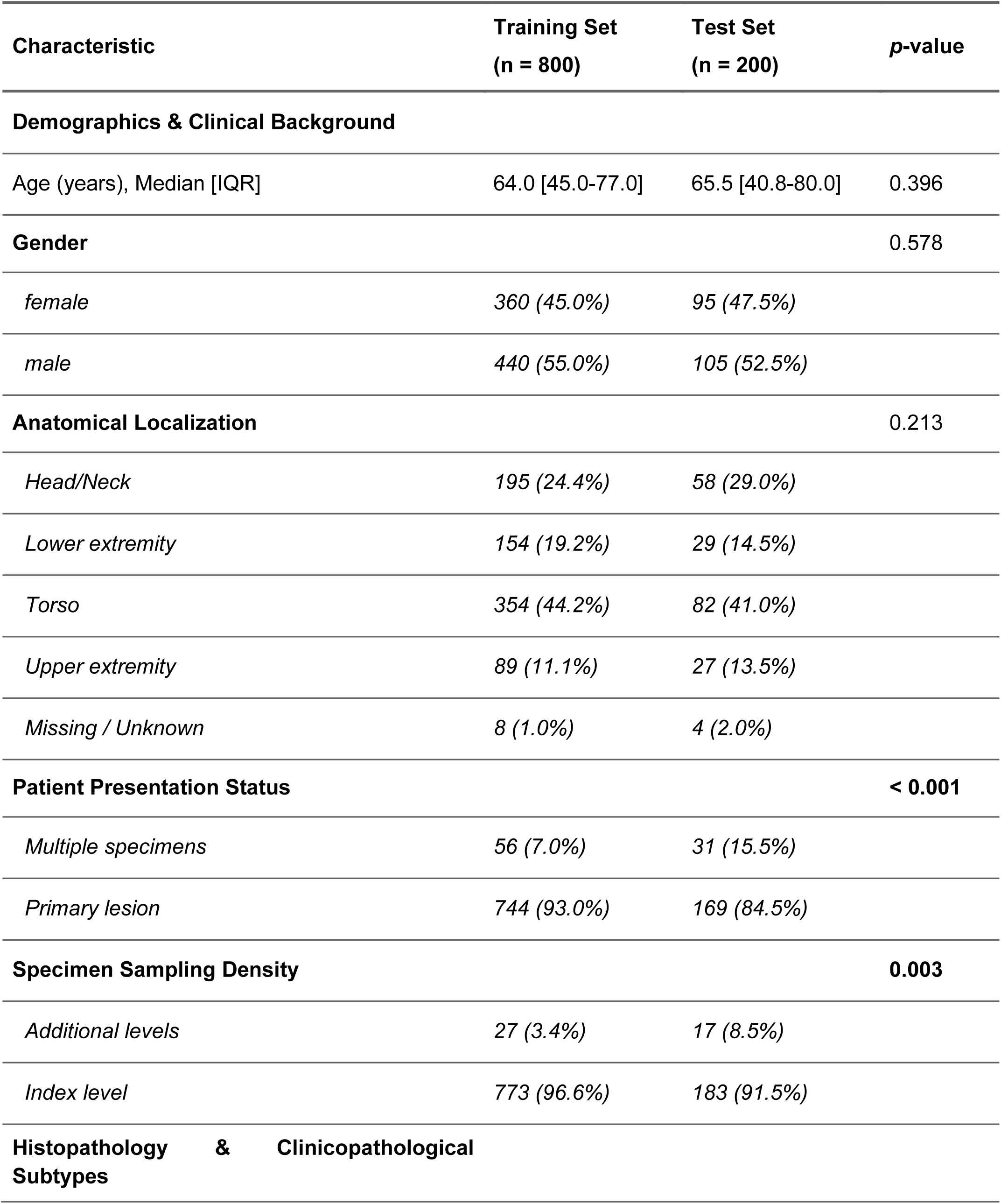
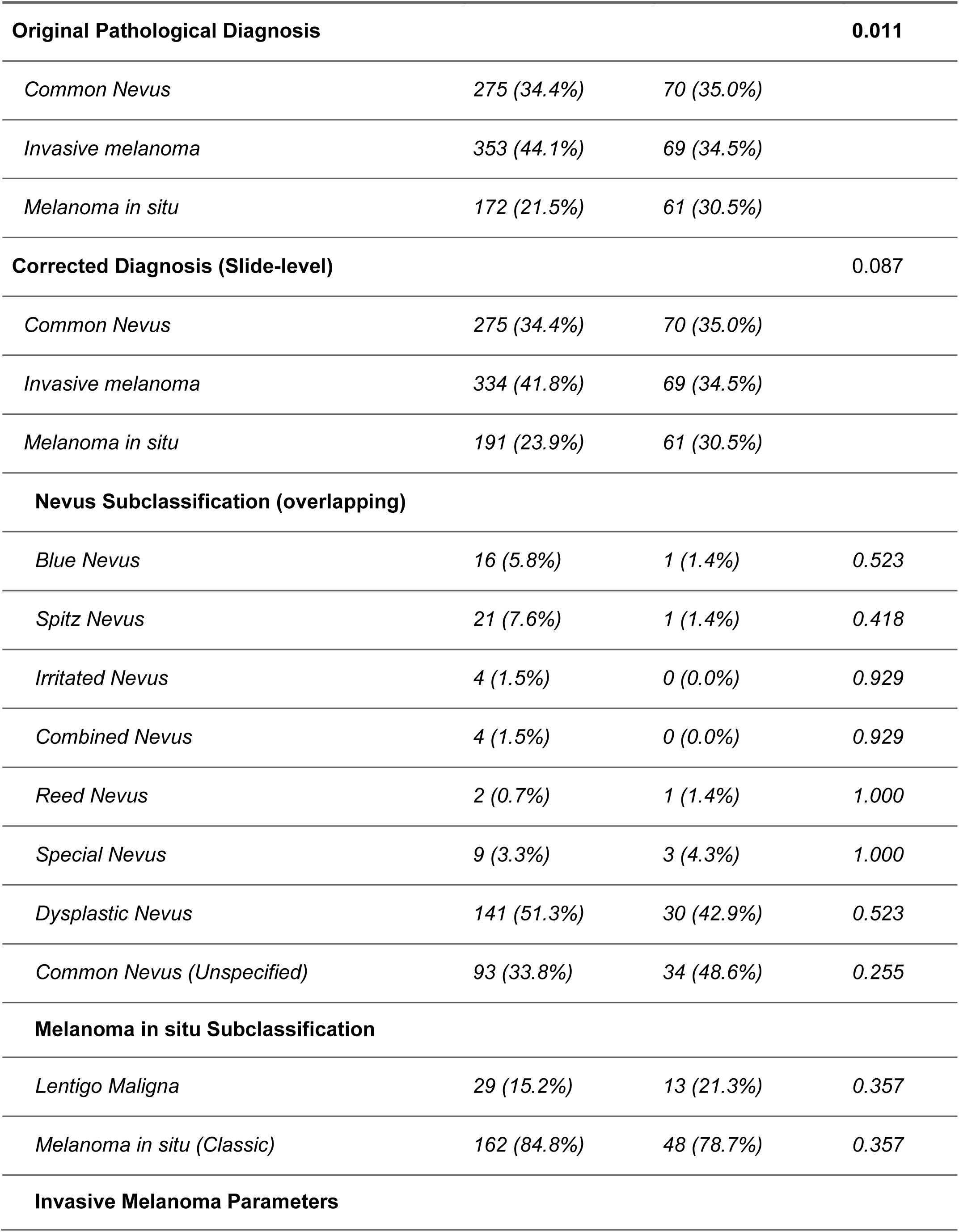
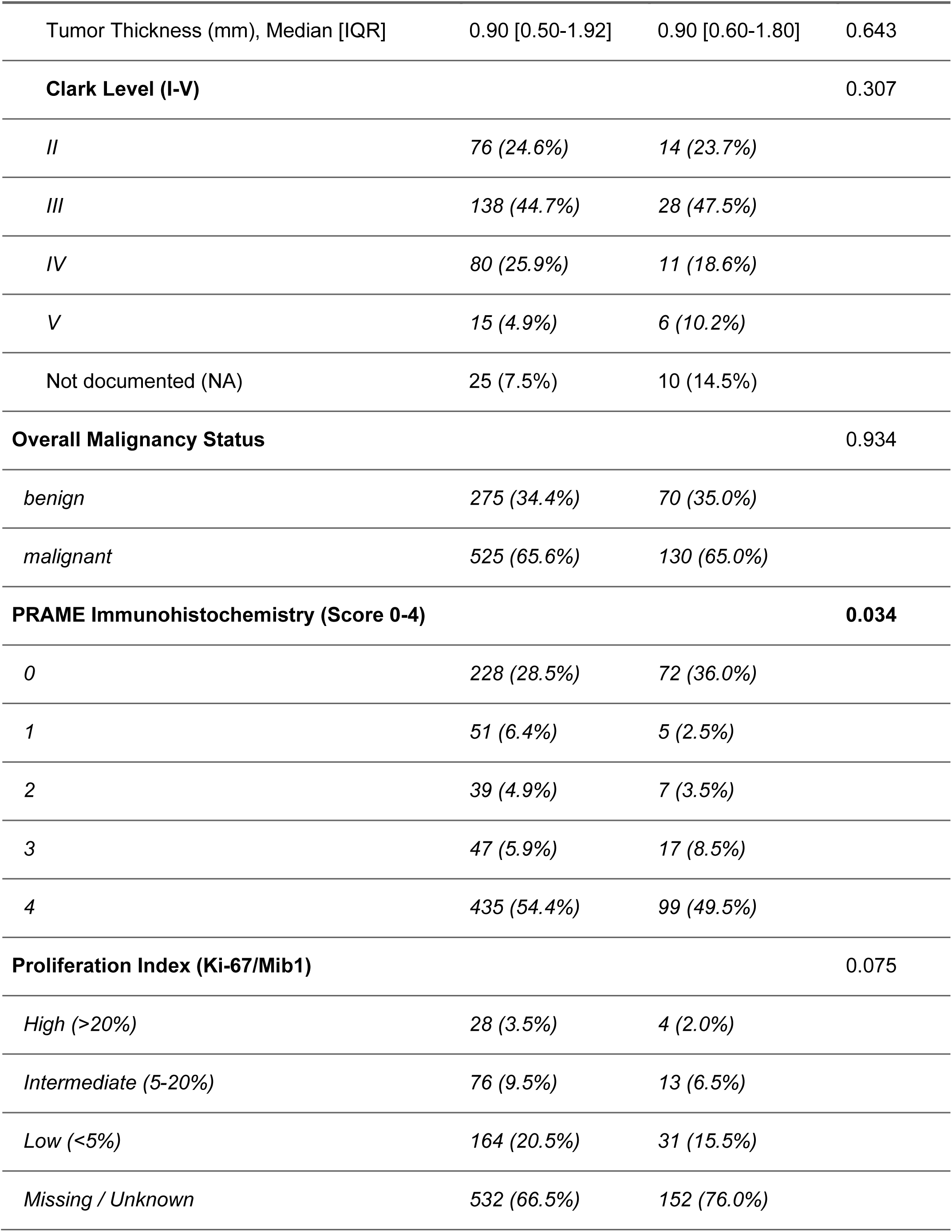
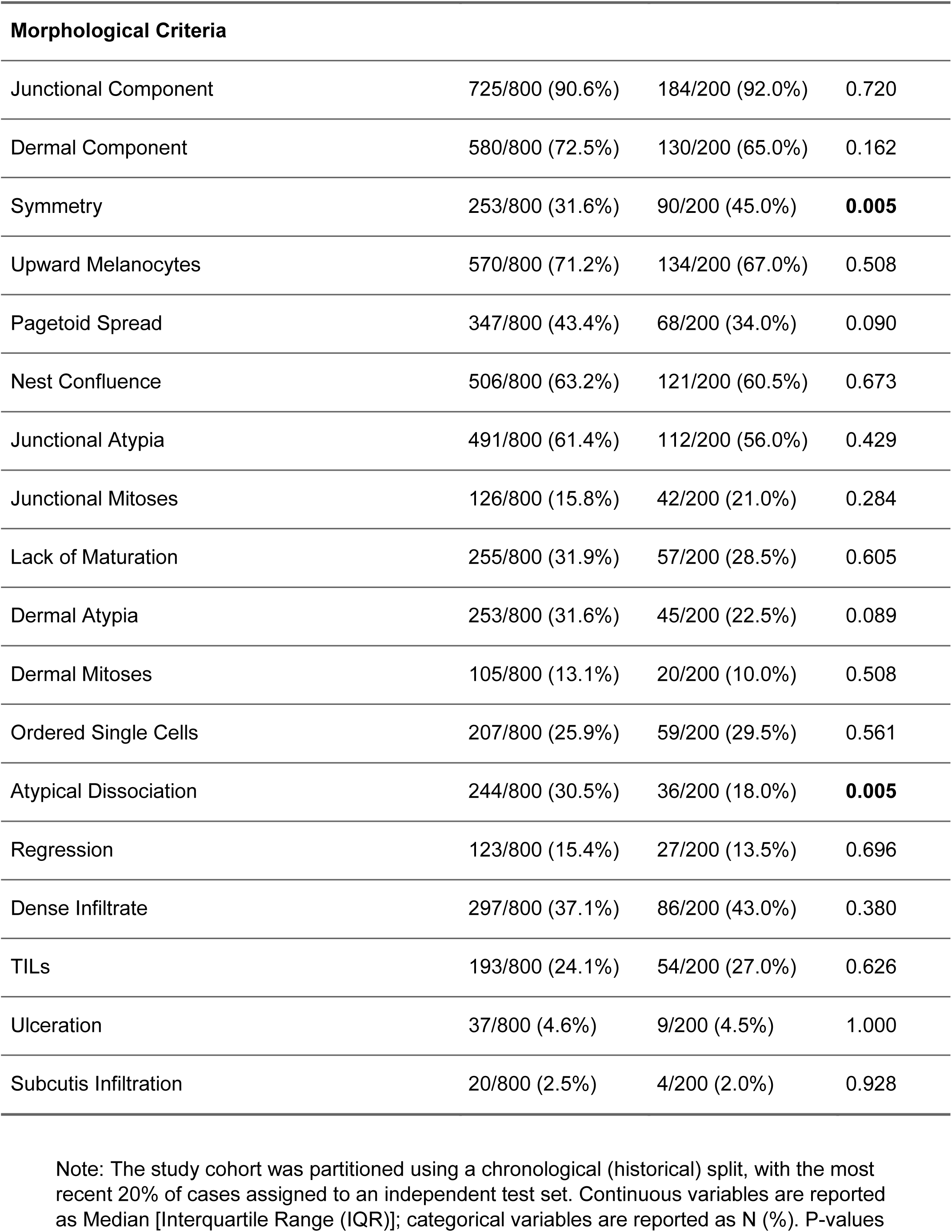
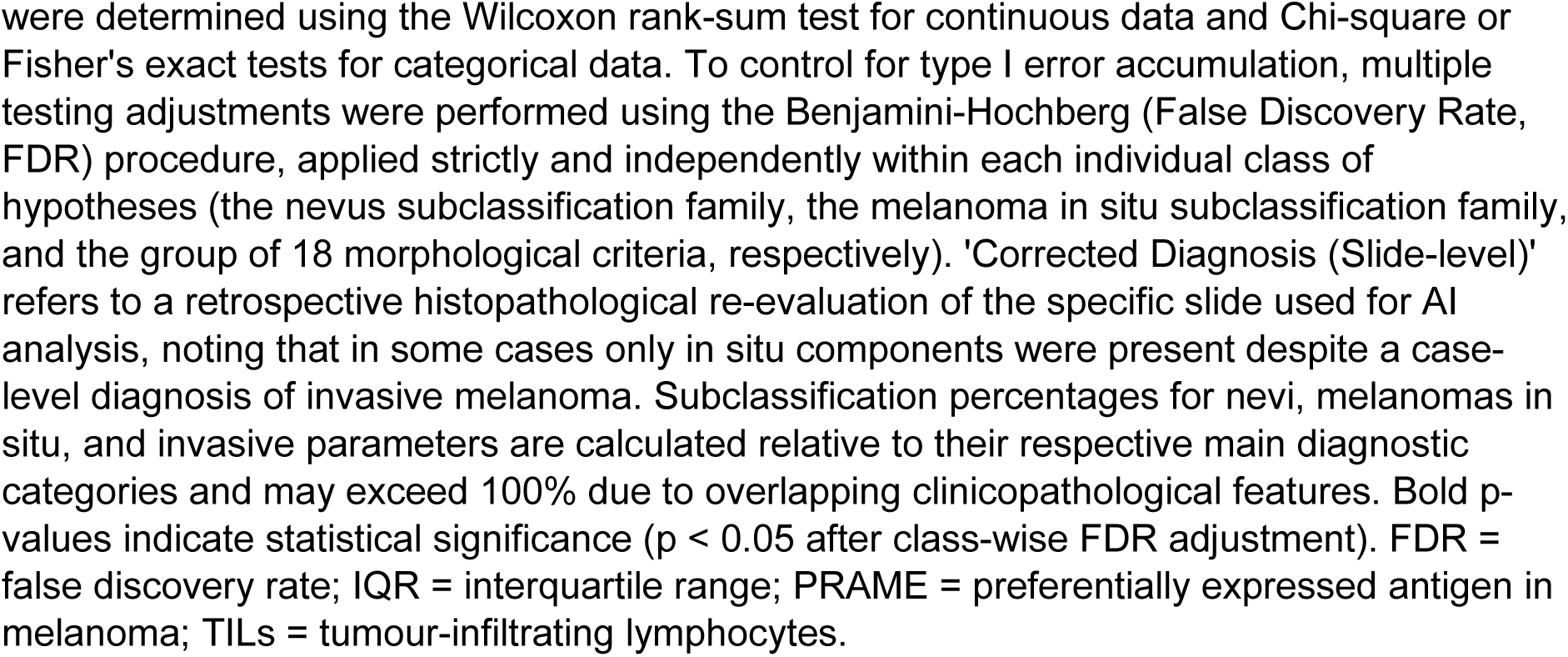
Baseline characteristics and morphological criteria of the study cohort.

| Characteristic | Training Set<br>(n = 800) | Test Set<br>(n = 200) | *p*-value |
|---|---|---|---|
| **Demographics & Clinical Background** |  |  |  |
| Age (years), Median [IQR] | 64.0 [45.0-77.0] | 65.5 [40.8-80.0] | 0.396 |
| **Gender** |  |  | 0.578 |
| *female* | *360 (45.0%)* | *95 (47.5%)* |  |
| *male* | *440 (55.0%)* | *105 (52.5%)* |  |
| **Anatomical Localization** |  |  | 0.213 |
| *Head/Neck* | *195 (24.4%)* | *58 (29.0%)* |  |
| *Lower extremity* | *154 (19.2%)* | *29 (14.5%)* |  |
| *Torso* | *354 (44.2%)* | *82 (41.0%)* |  |
| *Upper extremity* | *89 (11.1%)* | *27 (13.5%)* |  |
| *Missing / Unknown* | *8 (1.0%)* | *4 (2.0%)* |  |
| **Patient Presentation Status** |  |  | **< 0.001** |
| *Multiple specimens* | *56 (7.0%)* | *31 (15.5%)* |  |
| *Primary lesion* | *744 (93.0%)* | *169 (84.5%)* |  |
| **Specimen Sampling Density** |  |  | **0.003** |
| *Additional levels* | *27 (3.4%)* | *17 (8.5%)* |  |
| *Index level* | *773 (96.6%)* | *183 (91.5%)* |  |
| **Histopathology & Clinicopathological Subtypes** |  |  |  |
| **Original Pathological Diagnosis** |  |  | **0.011** |
| *Common Nevus* | *275 (34.4%)* | *70 (35.0%)* |  |
| *Invasive melanoma* | *353 (44.1%)* | *69 (34.5%)* |  |
| *Melanoma in situ* | *172 (21.5%)* | *61 (30.5%)* |  |
| **Corrected Diagnosis (Slide-level)** |  |  | 0.087 |
| *Common Nevus* | *275 (34.4%)* | *70 (35.0%)* |  |
| *Invasive melanoma* | *334 (41.8%)* | *69 (34.5%)* |  |
| *Melanoma in situ* | *191 (23.9%)* | *61 (30.5%)* |  |
| **Nevus Subclassification (overlapping)** |  |  |  |
| *Blue Nevus* | *16 (5.8%)* | *1 (1.4%)* | *0.523* |
| *Spitz Nevus* | *21 (7.6%)* | *1 (1.4%)* | *0.418* |
| *Irritated Nevus* | *4 (1.5%)* | *0 (0.0%)* | *0.929* |
| *Combined Nevus* | *4 (1.5%)* | *0 (0.0%)* | *0.929* |
| *Reed Nevus* | *2 (0.7%)* | *1 (1.4%)* | *1.000* |
| *Special Nevus* | *9 (3.3%)* | *3 (4.3%)* | *1.000* |
| *Dysplastic Nevus* | *141 (51.3%)* | *30 (42.9%)* | *0.523* |
| *Common Nevus (Unspecified)* | *93 (33.8%)* | *34 (48.6%)* | *0.255* |
| **Melanoma in situ Subclassification** |  |  |  |
| *Lentigo Maligna* | *29 (15.2%)* | *13 (21.3%)* | *0.357* |
| *Melanoma in situ (Classic)* | *162 (84.8%)* | *48 (78.7%)* | *0.357* |
| **Invasive Melanoma Parameters** |  |  |  |
| Tumor Thickness (mm), Median [IQR] | 0.90 [0.50-1.92] | 0.90 [0.60-1.80] | 0.643 |
| **Clark Level (I-V)** |  |  | 0.307 |
| *II* | *76 (24.6%)* | *14 (23.7%)* |  |
| *III* | *138 (44.7%)* | *28 (47.5%)* |  |
| *IV* | *80 (25.9%)* | *11 (18.6%)* |  |
| *V* | *15 (4.9%)* | *6 (10.2%)* |  |
| Not documented (NA) | 25 (7.5%) | 10 (14.5%) |  |
| **Overall Malignancy Status** |  |  | 0.934 |
| *benign* | *275 (34.4%)* | *70 (35.0%)* |  |
| *malignant* | *525 (65.6%)* | *130 (65.0%)* |  |
| **PRAME Immunohistochemistry (Score 0-4)** |  |  | **0.034** |
| *0* | *228 (28.5%)* | *72 (36.0%)* |  |
| *1* | *51 (6.4%)* | *5 (2.5%)* |  |
| *2* | *39 (4.9%)* | *7 (3.5%)* |  |
| *3* | *47 (5.9%)* | *17 (8.5%)* |  |
| *4* | *435 (54.4%)* | *99 (49.5%)* |  |
| **Proliferation Index (Ki-67/Mib1)** |  |  | 0.075 |
| *High (>20%)* | *28 (3.5%)* | *4 (2.0%)* |  |
| *Intermediate (5-20%)* | *76 (9.5%)* | *13 (6.5%)* |  |
| *Low (<5%)* | *164 (20.5%)* | *31 (15.5%)* |  |
| *Missing / Unknown* | *532 (66.5%)* | *152 (76.0%)* |  |
| **Morphological Criteria** |  |  |  |
| Junctional Component | 725/800 (90.6%) | 184/200 (92.0%) | 0.720 |
| Dermal Component | 580/800 (72.5%) | 130/200 (65.0%) | 0.162 |
| Symmetry | 253/800 (31.6%) | 90/200 (45.0%) | **0.005** |
| Upward Melanocytes | 570/800 (71.2%) | 134/200 (67.0%) | 0.508 |
| Pagetoid Spread | 347/800 (43.4%) | 68/200 (34.0%) | 0.090 |
| Nest Confluence | 506/800 (63.2%) | 121/200 (60.5%) | 0.673 |
| Junctional Atypia | 491/800 (61.4%) | 112/200 (56.0%) | 0.429 |
| Junctional Mitoses | 126/800 (15.8%) | 42/200 (21.0%) | 0.284 |
| Lack of Maturation | 255/800 (31.9%) | 57/200 (28.5%) | 0.605 |
| Dermal Atypia | 253/800 (31.6%) | 45/200 (22.5%) | 0.089 |
| Dermal Mitoses | 105/800 (13.1%) | 20/200 (10.0%) | 0.508 |
| Ordered Single Cells | 207/800 (25.9%) | 59/200 (29.5%) | 0.561 |
| Atypical Dissociation | 244/800 (30.5%) | 36/200 (18.0%) | **0.005** |
| Regression | 123/800 (15.4%) | 27/200 (13.5%) | 0.696 |
| Dense Infiltrate | 297/800 (37.1%) | 86/200 (43.0%) | 0.380 |
| TILs | 193/800 (24.1%) | 54/200 (27.0%) | 0.626 |
| Ulceration | 37/800 (4.6%) | 9/200 (4.5%) | 1.000 |
| Subcutis Infiltration | 20/800 (2.5%) | 4/200 (2.0%) | 0.928 |

Note: The study cohort was partitioned using a chronological (historical) split, with the most recent 20% of cases assigned to an independent test set. Continuous variables are reported as Median [Interquartile Range (IQR)]; categorical variables are reported as N (%). P-values were determined using the Wilcoxon rank-sum test for continuous data and Chi-square or Fisher's exact tests for categorical data. To control for type I error accumulation, multiple testing adjustments were performed using the Benjamini-Hochberg (False Discovery Rate, FDR) procedure, applied strictly and independently within each individual class of hypotheses (the nevus subclassification family, the melanoma in situ subclassification family, and the group of 18 morphological criteria, respectively). 'Corrected Diagnosis (Slide-level)' refers to a retrospective histopathological re-evaluation of the specific slide used for AI analysis, noting that in some cases only in situ components were present despite a case-level diagnosis of invasive melanoma. Subclassification percentages for nevi, melanomas in situ, and invasive parameters are calculated relative to their respective main diagnostic categories and may exceed 100% due to overlapping clinicopathological features. Bold p-values indicate statistical significance (p < 0.05 after class-wise FDR adjustment). FDR = false discovery rate; IQR = interquartile range; PRAME = preferentially expressed antigen in melanoma; TILs = tumour-infiltrating lymphocytes.

Standardized definitions for each morphological feature and scoring protocols for PRAME and Ki-67 are documented in the Supplementary Criteria Catalog (Supp. Material S2). Blinded to clinical history, the investigator recorded regression strictly as horizontal fibrosis and neovascularization.

### Deep Learning Pipeline and AI Analysis

Computational analysis used the open-source STAMP framework. Digitized WSIs underwent automated tissue segmentation to exclude background, and tissue regions were divided into non-overlapping tiles of 224×224 pixels (256×256 µm; 1.14 µm/pixel). Feature embeddings were extracted using the Virchow2 foundation model, mapping each tile to a 2,560-dimensional vector. Slide-level aggregation used an attention-based transformer model to train separate classifiers for binary malignancy, all 18 morphological criteria, and multiclass immunohistochemical markers (PRAME and Ki-67), evaluated via a one-versus-rest (OvR) approach. Complete hyperparameter specifications are provided in the GitHub repository.

### Evaluation and Validation Strategy

The dataset was partitioned chronologically, allocating the earliest 80% of cases (n=800) to the training set and the most recent 20% (n=200) to an independent test set. To prevent data leakage, allocation was performed at the patient level, assigning all lesions and sections from a single patient to the same partition. Model optimization used 5-fold cross-validation within the training cohort on NVIDIA CUDA-enabled GPUs, with final evaluation on the independent test set.

### Statistical Analysis

Statistical analyses were performed in R (version 4.4.3; tidyverse, pROC, PRROC, broom). Baseline comparisons used the Wilcoxon rank-sum test for continuous variables and Pearson’s chi-square or Fisher’s exact test for categorical variables. Model performance was quantified using the area under the receiver operating characteristic curve (AUROC; 95% CIs via DeLong’s method) and precision-recall curve (AUPRC), applying an OvR approach for multiclass endpoints.

Continuous AI probability scores for histological features were calibrated on the training cohort using logistic regression to establish operational decision thresholds. Concordance between calibrated AI predictions and the expert reference standard was evaluated using confusion matrices.

To evaluate predictor associations and compare diagnostic modalities, logistic regression models were fitted across three stages: the standalone slide-level AI malignancy model, a multivariable morphology model based on the 13 core features, and a multivariable logistic integration framework (’Triple Model’) combining the slide-level AI malignancy score, the 13 AI-derived morphological probabilities, and the clinical PRAME score (0–4). Predictor associations were quantified as odds ratios (ORs) with 95% CIs and p-values, including a pre-specified contrast comparing PRAME score 4 versus 0. All tests were two-sided (p<0.05).

## Results

### Study Population and Baseline Characteristics

The cohort comprised 1,000 primary cutaneous melanocytic lesions, split chronologically into a training cohort (n = 800) and an independent test cohort (n = 200; Figure 1, Table 1). In the training cohort (median age 64.0 years, IQR 45.0–77.0; 55.0% male), 525 lesions (65.6%) were malignant (353 invasive melanomas, 172 melanomas in situ) and 275 (34.4%) were benign nevi (Table 1). PRAME immunohistochemistry displayed a bimodal distribution, predominantly scoring 0 (28.5%) or 4 (54.4%), with intermediate scores (1–3) in 17.2% of cases.

The chronological split revealed diagnostic variations between cohorts (Table 1). While the proportion of benign nevi remained equivalent (35.0% vs. 34.4%), the independent test set displayed more melanomas in situ (30.5% vs. 21.5%) and fewer invasive melanomas (34.5% vs. 44.1%, p = 0.011; Table 1).

Histopathological feature evaluation also revealed compositional variations (Table 1). Lesional symmetry was more frequent in the test set (45.0% vs. 31.6%, p=0.005), whereas atypical dissociation was less prevalent (18.0% vs. 30.5%, p=0.005). Other criteria, including pagetoid spread, dermal atypia, and dermal mitoses, did not differ significantly (Table 1, Supplementary Table S1). Categorical PRAME expression also differed (p=0.034), with more score 0 (36.0% vs. 28.5%) and slightly fewer score 4 lesions (49.5% vs. 54.4%) in the test set (Table 1).

### Diagnostic Performance of the Slide-Level Malignancy Model

The slide-level AI malignancy model demonstrated high standalone diagnostic performance (Figure 2A, 2B). During 5-fold cross-validation on the training set (n = 800), the model achieved an AUROC of 0.925 (95% CI 0.907–0.943) and an AUPRC of 0.960. On the independent test set (n = 200), it achieved an AUROC of 0.970 (95% CI 0.951–0.989) and an AUPRC of 0.984 (Figure 2A, 2B), maintaining a precision of 1.00 up to a recall threshold of approximately 0.65 and identifying a majority of malignant cases without false-positive classifications (Figure 2B).

**Figure 2:**
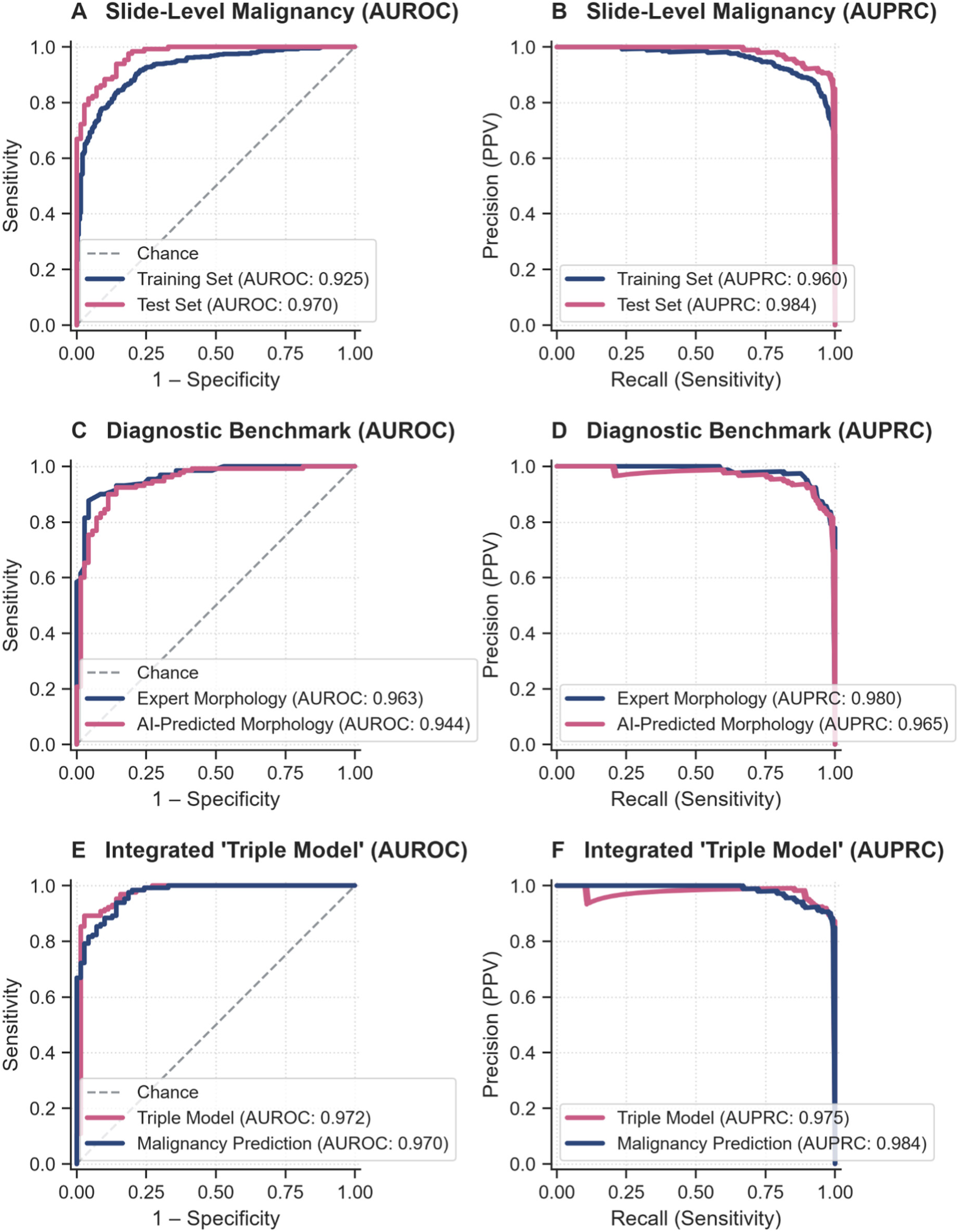
Diagnostic performance, morphological benchmarking, and multivariable integration of the foundation model. (A) AUROC and (B) AUPRC for the slide-level malignancy classifier across the 5-fold cross-validated training cohort (80% of cases) and the independent test cohort (20%). (C) AUROC and (D) AUPRC for the diagnostic benchmark evaluating multivariable models driven by expert-defined versus AI-predicted morphological features. (E) AUROC and (F) AUPRC comparing the integrated Triple Model against the slide-level malignancy classifier. AI = artificial intelligence. AUPRC = area under the precision-recall curve. AUROC = area under the receiver operating characteristic curve.

### Prediction of Histological Features and Immunohistochemical Markers

Separate slide-level classifiers were evaluated across the core panel of 13 morphological features, routine immunohistochemical markers (PRAME and Ki-67), and primary diagnostic categories (benign nevi, melanoma in situ, and invasive melanoma; Figure 3A; Supplementary Table S2).

**Figure 3:**
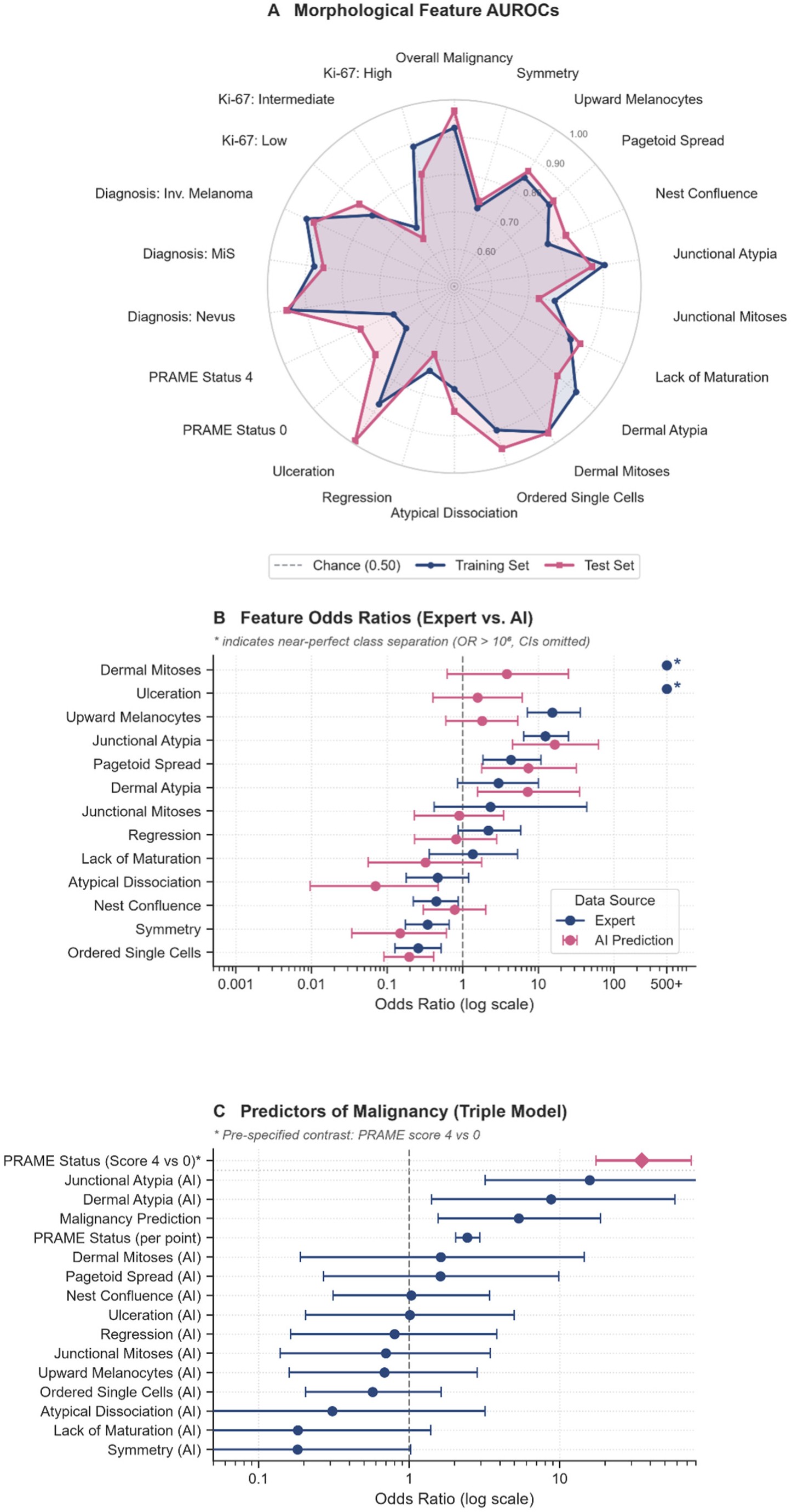
Feature-level diagnostic performance and multivariable predictors of malignancy. (A) Radar plot detailing AUROCs for AI predictions of morphological criteria, PRAME status, Ki-67 categories, and overall malignancy across the training and test cohorts. (B) Forest plot comparing multivariable odds ratios of morphological features between expert annotation and AI prediction; asterisks indicate extreme estimates resulting from near-perfect class separation. (C) Forest plot of multivariable odds ratios for predictors within the integrated Triple Model; the asterisk highlights the pre-specified contrast of PRAME score 4 versus 0. Error bars represent 95% confidence intervals. AI = artificial intelligence. AUROC = area under the receiver operating characteristic curve. OR = odds ratio. PRAME = preferentially expressed antigen in melanoma.

Morphological models demonstrated robust predictive performance across classic histological criteria in 5-fold cross-validation and independent testing (Figure 3A; Supplementary Table S2). Dermal mitoses showed the highest discriminatory consistency (training / test AUROC: 0.964 / 0.966), followed by ordered single cells (0.901 / 0.952) and junctional atypia (0.906 / 0.872), while ulceration achieved the highest test-set performance (0.875 / 0.991; Supplementary Table S2). Conversely, lower discrimination was observed for symmetry (0.719/ 0.737), regression (0.735 / 0.690), and junctional mitoses (0.772 / 0.730), reflecting moderate concordance with expert annotations (Supplementary Table S3).

Predicting immunohistochemical markers yielded lower discrimination than primary morphological criteria (Figure 3A; Supplementary Table S2). For PRAME expression, classifiers achieved training / test AUROCs of 0.671 / 0.780 for score 0 and 0.679 / 0.776 for score 4, while predicting a high Ki-67 proliferation index yielded 0.890 / 0.812 (Figure 3A; Supplementary Table S2).

Across the feature spectrum, the slide-level malignancy classifier exceeded the discrimination of all individual morphological and immunohistochemical targets, except for ulceration in the independent test set (Figure 3A; Supplementary Table S2).

### Benchmarking Morphological Models Against Expert Annotations

In multivariable logistic regression models trained on the 13 core morphological criteria, the expert-annotated model achieved an AUROC of 0.963 (95% CI 0.940–0.986) in the independent test set, compared to 0.944 (95% CI 0.910–0.977) for AI-predicted features, with overlapping 95% confidence intervals (Figure 2C, 2D).

Multivariable analysis demonstrated both concordance and divergence in feature weighting (Figure 3B; Supplementary Table S4). In the expert-annotated model, dermal mitoses and ulceration caused quasi-complete class separation, precluding stable parameter estimation. Among evaluable features, both models identified junctional atypia as the strongest independent predictor of malignancy (expert OR 12.42, AI OR 16.54; both p < 0.001), while ordered single cells consistently indicated benignity (expert OR 0.26 vs. AI OR 0.19; both p < 0.001). Key divergences emerged for upward-migrating melanocytes, predictive only in the expert model (OR 15.35, p < 0.001 vs. OR 1.80, p = 0.292), and atypical dissociation, associated with benignity only in the AI model (OR 0.07, p = 0.007).

### Integration of Malignancy Prediction, Morphology, and PRAME Status (Triple Model)

Finally, the integrated model was evaluated, incorporating clinical PRAME status alongside the 13 AI-predicted morphological criteria and the slide-level malignancy prediction score. In the independent test set, the Triple Model achieved an AUROC of 0.972 (95% CI 0.944–0.999) and an AUPRC of 0.975, closely comparable to the standalone malignancy classifier (AUROC 0.970; AUPRC 0.984) with broadly overlapping confidence intervals (Figure 2E, 2F; Table 2).

**Table 2:** Odds Ratios for the Triple Model (including PRAME Score 4 vs 0 comparison).

| **Feature** | **aOR [95% CI]** | **p-value** |
|---|---|---|
| **PRAME Status (Score 4 vs 0)\*** | **35.04 [17.39 - 74.62]** | **< .001** |
| Junctional Atypia (AI) | 15.84 [3.20 - 82.44] | **< .001** |
| Dermal Atypia (AI) | 8.77 [1.42 - 58.20] | **0.022** |
| Malignancy Prediction | 5.34 [1.56 - 18.68] | **0.008** |
| PRAME Status (per point) | 2.43 [2.04 - 2.94] | **< .001** |
| Dermal Mitoses (AI) | 1.62 [0.19 - 14.60] | 0.662 |
| Pagetoid Spread (AI) | 1.62 [0.27 - 9.85] | 0.598 |
| Nest Confluence (AI) | 1.03 [0.31 - 3.43] | 0.956 |
| Ulceration (AI) | 1.01 [0.20 - 4.98] | 0.990 |
| Regression (AI) | 0.80 [0.16 - 3.82] | 0.779 |
| Junctional Mitoses (AI) | 0.70 [0.14 - 3.45] | 0.661 |
| Upward Melanocytes (AI) | 0.68 [0.16 - 2.83] | 0.605 |
| Ordered Single Cells (AI) | 0.57 [0.20 - 1.63] | 0.292 |
| Atypical Dissociation (AI) | 0.31 [0.03 - 3.20] | 0.326 |
| Lack of Maturation (AI) | 0.18 [0.02 - 1.39] | 0.104 |
| Symmetry (AI) | 0.18 [0.03 - 1.03] | 0.056 |

Note: Multivariable logistic regression of predictors in the Triple Model. The synthetic 'PRAME Score 4 vs 0' row estimates the cumulative effect size for a maximum PRAME score increase. Bold p-values indicate statistical significance (p < 0.05). AI = artificial intelligence; aOR = adjusted odds ratio; CI = confidence interval; PRAME = preferentially expressed antigen in melanoma.

In multivariable logistic regression, PRAME status was significantly associated with malignancy (OR 2.43 per incremental point, p < 0.001), corresponding to an OR of 35.04 comparing maximal expression against complete absence (score 4 versus 0, p < 0.001; Figure 3C; Table 2). Following PRAME, junctional atypia (OR 15.84, p < 0.001), dermal atypia (OR 8.77, p = 0.022), and the slide-level malignancy classifier (OR 5.34, p = 0.008) emerged as the strongest independent predictors of malignancy.

## Discussion

### Cohort Selection and Temporal Validation

Defining the cohort by clinical requests for PRAME immunohistochemistry enriched the dataset for lesions with diagnostic ambiguity and high inter-observer discordance ^5,6^. Applying a chronological rather than random train-test split introduced natural temporal domain shifts in tissue processing, scanner calibration, and disease prevalence, closely reflecting prospective deployment. Despite compositional differences, particularly a higher proportion of melanoma in situ in the test set, diagnostic performance remained stable, indicating that extracted representations capture invariant morphological features rather than temporal laboratory artifacts.

### Slide-Level Malignancy Classification

The slide-level malignancy classifier demonstrated high standalone accuracy. While test-cohort performance slightly exceeded internal cross-validation, this difference likely reflects cohort composition and sampling variability rather than systematic shifts in diagnostic practice.

Clinically, by maintaining complete precision up to a substantial recall threshold, the classifier identified two-thirds of malignant lesions without false-positive diagnoses in the test cohort. In routine pathology, this profile could support automated triage, prioritizing high-confidence cases before expert review, although prospective validation with predefined operating thresholds remains required to confirm clinical utility.

This performance in borderline melanocytic proliferations marks a clear advance over early-generation machine learning approaches. Whereas multi-center meta-analyses of task-specific CNNs reported pooled concordance indices around 0.87 ^34^, large-scale foundation models such as Virchow2 ^20^ utilize self-supervised pre-training across millions of WSIs to overcome representational bottlenecks and enable clinical-grade decision support.

### Prediction of Morphological Criteria and the Immunohistochemical Gap

Evaluating the 13 histological criteria confirmed that foundation model representations support predicting established histopathological features ^28^. Structural criteria such as ulceration, dermal mitoses, and ordered single cells were predicted with high accuracy, reflecting dermatopathological domain knowledge. In contrast, discrete cytomorphological criteria proved more challenging to resolve, reflecting the difficulty of capturing high-power nuclear details spanning few pixels within tile embeddings, where localized signals are diluted during slide-level attention pooling.

In multivariable regression, models driven by automated feature predictions achieved diagnostic performance comparable to expert human annotations. Beyond classification accuracy, translating continuous slide embeddings into discrete histological criteria aligns deep-learning representations with conventional pathological reasoning, establishing a transparent foundation for human-in-the-loop workflows ^24–27,35^.

A performance drop emerged when predicting immunohistochemical status directly from standard H&E morphology, with classifiers for PRAME expression or Ki-67 proliferation performing substantially worse than morphological predictors. While computational models can predict specific molecular alterations from routine histology across cancers ^29^, PRAME expression in diagnostically challenging melanocytic lesions cannot be reliably predicted from H&E alone. In this borderline setting, targeted immunohistochemistry provides non-redundant biological information not fully encoded within standard histological patterns.

### The Triple Model with Clinical PRAME Integration

Because immunohistochemical status cannot be reliably inferred from routine histology alone, the Triple Model integrated clinical PRAME scoring with AI-predicted morphological features and the slide-level malignancy classifier, preserving high predictive performance comparable to the standalone classifier.

Within this multivariable framework, clinical PRAME status emerged as a strongly associated independent predictor alongside automated predictions of junctional atypia, dermal atypia, and the slide-level classifier. Thus, automated morphology and immunohistochemistry provide complementary, non-redundant diagnostic value.

While proof-of-concept studies explored digital PRAME quantification in small cohorts ^30^, our study demonstrates across 1,000 lesions that integrating this biomarker directly with an H&E foundation model provides a practical framework for clinical decision support. The Triple Model highlights that the greatest utility of computational dermatopathology lies in combining physical biomarkers with automated morphological analysis rather than predicting immunohistochemistry purely from H&E ^32,33^.

### Limitations

This study has several limitations. First, while chronological splitting provides rigorous temporal validation, the retrospective single-center design necessitates external validation across diverse scanning hardware and independent cohorts. Second, annotation by a single pathologist ensures high internal consistency but does not capture inter-observer variability, underscoring the need for multi-reader validation. Third, the integrated Triple Model did not substantially improve discrimination over the standalone slide-level classifier, indicating that the incremental clinical benefit of model integration requires prospective evaluation. Finally, restricting the cohort to cases with clinically requested PRAME staining introduces selection bias. Thus, our findings reflect a pre-filtered, challenging diagnostic spectrum rather than unselected routine biopsies.

## Conclusion

This study demonstrates that histopathology foundation models achieve robust diagnostic discrimination in identifying malignant melanoma within clinically challenging, borderline lesions. Automated profiling of discrete histological criteria translates continuous image embeddings into transparent features aligned with human expertise. However, key molecular biomarkers cannot be replaced by routine morphology alone. By uniting standard H&E features with clinical PRAME scoring, the integrated Triple Model combines high diagnostic accuracy with interpretability for computer-assisted decision support in dermatopathology.

## Declaration of Generative AI and AI-Assisted Technologies in the Writing Process

During the preparation of this manuscript, the authors utilized Gemini (Google) as an editorial and computational assistant. Specifically, the tool was employed to expand author-generated notes and bulleted drafts into continuous prose, refine syntax, condense text to meet journal constraints, and assist in drafting and optimizing R, Bash, and Python scripts utilized for data processing and statistical workflows. All scientific concepts, study designs, analytical pipelines, and data interpretations originated entirely from the authors. Following AI-assisted drafting and script generation, the authors rigorously reviewed, tested, verified, and refined all code and textual content to ensure complete computational reproducibility, factual integrity, and scientific accuracy. The authors take full responsibility for the final text, data analyses, and overall integrity of the published work.

## Ethical Approval Statement

The study was conducted in accordance with the Declaration of Helsinki and received formal approval from the Ethics Committee at TU Dresden (decision reference: BO-EK-172042024).

## Ethics Statement

The requirement for informed patient consent was waived in accordance with Section 29 of the Saxon Hospital Act (SächsKHG) for retrospective analysis of routinely collected clinical data. No identifiable individual patient data or identifiable photographs are included in this study.

## Data Availability Statement

The aggregated and anonymized prediction data, R analysis scripts, and model configurations required to reproduce the findings of this study are openly available in the GitHub repository at https://github.com/ffgellrich/ai-prame-dermatopathology and permanently archived on Zenodo (https://doi.org/10.5281/zenodo.24220711). Due to institutional data privacy regulations regarding sensitive patient health records, raw whole-slide images (WSIs) and unanonymized clinical metadata cannot be made publicly available.

## Author Contributions

**Conceptualization:** Frank F. Gellrich, Claudia Günther, Jakob N. Kather, and Narmin Ghaffari Laleh.

**Data curation:** Frank F. Gellrich and Alaska Kistner.

**Formal analysis:** Frank F. Gellrich, Alaska Kistner, Jakob N. Kather, and Narmin Ghaffari Laleh.

**Investigation:** Cosima Hufnagel, Jörg Laske, Daniela Aust, Gustavo Baretton, Nick Reidow, Mildred Sergon, and Narmin Ghaffari Laleh.

**Methodology:** Frank F. Gellrich, Jakob N. Kather, and Narmin Ghaffari Laleh.

**Project administration:** Frank F. Gellrich and Jakob N. Kather.

**Resources:** Frank F. Gellrich, Alaska Kistner, Sarah Hobelsberger, Stefan Beissert, Daniela Aust, Gustavo Baretton, Jakob N. Kather, and Narmin Ghaffari Laleh.

**Software:** Frank F. Gellrich, Alaska Kistner, Jakob N. Kather, and Narmin Ghaffari Laleh.

**Supervision:** Frank F. Gellrich, Jakob N. Kather, and Narmin Ghaffari Laleh.

**Validation:** Frank F. Gellrich, Alaska Kistner, Jakob N. Kather, and Narmin Ghaffari Laleh.

**Visualization:** Frank F. Gellrich and Narmin Ghaffari Laleh.

**Writing - original draft:** Frank F. Gellrich and Narmin Ghaffari Laleh.

**Writing - review & editing:** Frank F. Gellrich, Alaska Kistner, Claudia Günther, Cosima Hufnagel, Sarah Hobelsberger, Jörg Laske, Stefan Beissert, Daniela Aust, Gustavo Baretton, Nick Reidow, Mildred Sergon, Jakob N. Kather, and Narmin Ghaffari Laleh.

All authors critically reviewed, contributed to, and approved the final manuscript.

