## Supplementary Material for "Integrating Histopathology Foundation Models and PRAME Immunohistochemistry in Melanoma Diagnosis"

### Full Title

### Names of Authors

Gellrich, Frank Friedrich[1,2,3,*]

Kistner, Alaska[1]

Günther, Claudia[1,6]

Hufnagel, Cosima[1]

Hobelsberger, Sarah[1]

Laske, Jörg[1]

Beissert, Stefan[1]

Aust, Daniela[3]

Baretton, Gustavo[3]

Reidow, Nick[3]

Sergon, Mildred[3]

Kather, Jakob Nikolas[4,5,†]

Ghaffari Laleh, Narmin[4,†]

#### Footnotes

[*]Corresponding author: Dr. med. Frank Friedrich Gellrich

[†]These authors contributed equally

### Institutional Affiliations

[1]Department of Dermatology, Faculty of Medicine and University Hospital Carl Gustav Carus, Technische Universität Dresden, Dresden, Germany

[2]Skin Cancer Center at the University Cancer Center and National Center for Tumor Diseases Dresden, Dresden, Germany

[3]Institute of Pathology, Faculty of Medicine and University Hospital Carl Gustav Carus, Technische Universität Dresden, Dresden, Germany

[4]Else Kroener Fresenius Center for Digital Health, Technical University Dresden, Dresden, Germany

[5]Department of Medicine I, Faculty of Medicine and University Hospital Carl Gustav Carus, Technische Universität Dresden, Dresden, Germany

[6]Department of Dermatology, Faculty of Medicine and University Hospital Tübingen, Eberhard Karls University of Tübingen, Tübingen, Germany

### Supplementary Tables

#### Supp. Table S1: Calibrated Feature Frequencies (Training Set, CV80)

| Criterion | Attribute | GT Malignant | GT Benign | AI Malignant | AI Benign |
|---|---|---|---|---|---|
| Overall Malignancy | Present | 525/525 (100.0%) | 0/275 (0.0%) | 464/525 (88.4%) | 57/275 (20.7%) |
| Symmetry | Present | 80/525 (15.2%) | 173/275 (62.9%) | 26/525 (5.0%) | 109/275 (39.6%) |
| Upward Melanocytes | Present | 493/525 (93.9%) | 77/275 (28.0%) | 486/525 (92.6%) | 104/275 (37.8%) |
| Pagetoid Spread | Present | 338/525 (64.4%) | 9/275 (3.3%) | 351/525 (66.9%) | 15/275 (5.5%) |
| Nest Confluence | Present | 361/525 (68.8%) | 145/275 (52.7%) | 395/525 (75.2%) | 180/275 (65.5%) |
| Junctional Atypia | Present | 462/525 (88.0%) | 29/275 (10.5%) | 451/525 (85.9%) | 39/275 (14.2%) |
| Junctional Mitoses | Present | 125/525 (23.8%) | 1/275 (0.4%) | 0/525 (0.0%) | 0/275 (0.0%) |
| Lack of Maturation | Present | 241/525 (45.9%) | 14/275 (5.1%) | 202/525 (38.5%) | 15/275 (5.5%) |
| Dermal Atypia | Present | 240/525 (45.7%) | 13/275 (4.7%) | 246/525 (46.9%) | 18/275 (6.5%) |
| Dermal Mitoses | Present | 105/525 (20.0%) | 0/275 (0.0%) | 112/525 (21.3%) | 5/275 (1.8%) |
| Ordered Single Cells | Present | 45/525 (8.6%) | 162/275 (58.9%) | 48/525 (9.1%) | 167/275 (60.7%) |
| Atypical Dissociation | Present | 210/525 (40.0%) | 34/275 (12.4%) | 186/525 (35.4%) | 12/275 (4.4%) |
| Regression | Present | 106/525 (20.2%) | 17/275 (6.2%) | 3/525 (0.6%) | 0/275 (0.0%) |
| Ulceration | Present | 37/525 (7.0%) | 0/275 (0.0%) | 18/525 (3.4%) | 0/275 (0.0%) |

| Criterion | Attribute | GT Malignant | GT Benign | AI Malignant | AI Benign |
|---|---|---|---|---|---|
| Junctional Component | Present | 508/525 (96.8%) | 217/275 (78.9%) | 525/525 (100.0%) | 275/275 (100.0%) |
| Dermal Component | Present | 363/525 (69.1%) | 217/275 (78.9%) | 358/525 (68.2%) | 203/275 (73.8%) |
| Dense Lymphocytic Infiltrate | Present | 248/525 (47.2%) | 49/275 (17.8%) | 259/525 (49.3%) | 37/275 (13.5%) |
| Tumor-Infiltrating Lymphocytes (TILs) | Present | 170/525 (32.4%) | 23/275 (8.4%) | 175/525 (33.3%) | 10/275 (3.6%) |
| Subcutis Infiltration | Present | 15/525 (2.9%) | 5/275 (1.8%) | 0/525 (0.0%) | 0/275 (0.0%) |
| PRAME Status | Score 0 | 36/525 (6.9%) | 192/275 (69.8%) | 60/525 (11.4%) | 71/275 (25.8%) |
| | Score 1 | 21/525 (4.0%) | 30/275 (10.9%) | 44/525 (8.4%) | 34/275 (12.4%) |
| | Score 2 | 20/525 (3.8%) | 19/275 (6.9%) | 114/525 (21.7%) | 85/275 (30.9%) |
| | Score 3 | 32/525 (6.1%) | 15/275 (5.5%) | 88/525 (16.8%) | 38/275 (13.8%) |
| | Score 4 | 416/525 (79.2%) | 19/275 (6.9%) | 219/525 (41.7%) | 47/275 (17.1%) |
| Ki-67 Index (Mib1) | Low (<5%) | 67/170 (39.4%) | 97/98 (99.0%) | 49/170 (28.8%) | 73/98 (74.5%) |
| | Intermediate (5-20%) | 75/170 (44.1%) | 1/98 (1.0%) | 80/170 (47.1%) | 16/98 (16.3%) |
| | High (>20%) | 28/170 (16.5%) | 0/98 (0.0%) | 41/170 (24.1%) | 9/98 (9.2%) |

Note: AI binary predictions for morphological criteria were recalculated using logistic regression models calibrated on the training set (GT_Feature ~ AI_Feature_Score) to provide optimized frequency estimates beyond the standard 0.5 threshold. Malignant/Benign stratification is based on the Ground Truth diagnosis.

Supp. Table S2: Comprehensive AUC Performance Metrics across all diagnostic features.

| Feature | N (CV) | AUC CV [95% CI] | N (Deploy) | AUC Deploy [95% CI] |
|---|---|---|---|---|
| Overall Malignancy | 800 | 0.925 [0.907-0.943] | 200 | 0.970 [0.951-0.989] |
| Symmetry | 800 | 0.719 [0.681-0.757] | 200 | 0.737 [0.666-0.808] |
| Upward Melanocytes | 800 | 0.847 [0.818-0.877] | 200 | 0.867 [0.811-0.924] |
| Pagetoid Spread | 800 | 0.835 [0.808-0.863] | 200 | 0.850 [0.798-0.903] |
| Nest Confluence | 800 | 0.775 [0.742-0.809] | 200 | 0.828 [0.771-0.886] |
| Junctional Atypia | 800 | 0.906 [0.885-0.927] | 200 | 0.872 [0.825-0.920] |
| Junctional Mitoses | 800 | 0.772 [0.730-0.813] | 200 | 0.730 [0.641-0.818] |
| Lack of Maturation | 800 | 0.842 [0.813-0.871] | 200 | 0.870 [0.808-0.931] |
| Dermal Atypia | 800 | 0.931 [0.913-0.948] | 200 | 0.866 [0.793-0.939] |
| Dermal Mitoses | 800 | 0.964 [0.948-0.980] | 200 | 0.966 [0.938-0.995] |
| Ordered Single Cells | 800 | 0.901 [0.877-0.924] | 200 | 0.952 [0.923-0.982] |
| Atypical Dissociation | 800 | 0.775 [0.741-0.809] | 200 | 0.834 [0.752-0.917] |
| Regression | 800 | 0.735 [0.689-0.782] | 200 | 0.690 [0.590-0.789] |
| Ulceration | 800 | 0.875 [0.807-0.942] | 200 | 0.991 [0.980-1.000] |
| PRAME Status 0 | 800 | 0.671 [0.631-0.712] | 200 | 0.780 [0.713-0.846] |
| PRAME Status 4 | 800 | 0.679 [0.642-0.717] | 200 | 0.776 [0.711-0.840] |
| Diagnosis: Nevus | 800 | 0.946 [0.931-0.961] | 200 | 0.954 [0.928-0.979] |
| Diagnosis: MiS | 800 | 0.879 [0.853-0.906] | 200 | 0.854 [0.800-0.907] |
| Diagnosis: Inv. Melanoma | 800 | 0.935 [0.919-0.951] | 200 | 0.913 [0.872-0.954] |
| Ki-67: Low | 268 | 0.792 [0.738-0.845] | 48 | 0.837 [0.697-0.976] |
| Ki-67: Intermediate | 268 | 0.688 [0.619-0.757] | 48 | 0.653 [0.464-0.842] |
| Ki-67: High | 268 | 0.890 [0.824-0.957] | 48 | 0.812 [0.539-1.000] |

Note: Performance metrics (AUC) are stratified by Training Set (80%) and Independent Test Set (20%). For multiclass variables such as PRAME status, pathological diagnosis, and Ki-67 index, category-specific AUCs were calculated using a One-vs-Rest (OvR) approach against all other categories. Confidence Intervals (CI) were determined using the DeLong method. Abbreviations: AUC, Area Under the Receiver Operating Characteristic Curve; CI, Confidence Interval.

Supp. Table S3: Agreement Matrix (Training Set, N=800)

| Criterion | Agreement Both (TP) | Human only (FN) | AI only (FP) | Agreement Both (TN) | Total Human GT (+) | Total AI Pred (+) |
|---|---|---|---|---|---|---|
| **Overall Malignancy** | 464 (58.0%) | 61 (7.6%) | 57 (7.1%) | 218 (27.3%) | 525 (65.6%) | 521 (65.1%) |
| **Symmetry** | 79 (9.9%) | 174 (21.8%) | 56 (7.0%) | 491 (61.4%) | 253 (31.6%) | 135 (16.9%) |
| **Upward Melanocytes** | 500 (62.5%) | 70 (8.8%) | 90 (11.2%) | 140 (17.5%) | 570 (71.2%) | 590 (73.8%) |
| **Pagetoid Spread** | 262 (32.8%) | 85 (10.6%) | 104 (13.0%) | 349 (43.6%) | 347 (43.4%) | 366 (45.8%) |
| **Nest Confluence** | 433 (54.1%) | 73 (9.1%) | 142 (17.8%) | 152 (19.0%) | 506 (63.2%) | 575 (71.9%) |
| **Junctional Atypia** | 424 (53.0%) | 67 (8.4%) | 66 (8.2%) | 243 (30.4%) | 491 (61.4%) | 490 (61.3%) |
| **Junctional Mitoses** | 0 (0.0%) | 126 (15.8%) | 0 (0.0%) | 674 (84.2%) | 126 (15.8%) | 0 (0.0%) |
| **Lack of Maturation** | 156 (19.5%) | 99 (12.4%) | 61 (7.6%) | 484 (60.5%) | 255 (31.9%) | 217 (27.1%) |
| **Dermal Atypia** | 204 (25.5%) | 49 (6.1%) | 60 (7.5%) | 487 (60.9%) | 253 (31.6%) | 264 (33.0%) |
| **Dermal Mitoses** | 83 (10.4%) | 22 (2.8%) | 34 (4.2%) | 661 (82.6%) | 105 (13.1%) | 117 (14.6%) |
| **Ordered Single Cells** | 149 (18.6%) | 58 (7.2%) | 66 (8.2%) | 527 (65.9%) | 207 (25.9%) | 215 (26.9%) |
| **Atypical Dissociation** | 121 (15.1%) | 123 (15.4%) | 77 (9.6%) | 479 (59.9%) | 244 (30.5%) | 198 (24.8%) |
| **Regression** | 3 (0.4%) | 120 (15.0%) | 0 (0.0%) | 677 (84.6%) | 123 (15.4%) | 3 (0.4%) |
| **Ulceration** | 15 (1.9%) | 22 (2.8%) | 3 (0.4%) | 760 (95.0%) | 37 (4.6%) | 18 (2.2%) |
| **Junctional Component** | 725 (90.6%) | 0 (0.0%) | 75 (9.4%) | 0 (0.0%) | 725 (90.6%) | 800 (100.0%) |

| Criterion | Agreement Both (TP) | Human only (FN) | AI only (FP) | Agreement Both (TN) | Total Human GT (+) | Total AI Pred (+) |
|---|---|---|---|---|---|---|
| **Dermal Component** | 504 (63.0%) | 76 (9.5%) | 57 (7.1%) | 163 (20.4%) | 580 (72.5%) | 561 (70.1%) |
| **Dense Lymphocytic Infiltrate** | 220 (27.5%) | 77 (9.6%) | 76 (9.5%) | 427 (53.4%) | 297 (37.1%) | 296 (37.0%) |
| **Tumor-Infiltrating Lymphocytes** | 114 (14.2%) | 79 (9.9%) | 71 (8.9%) | 536 (67.0%) | 193 (24.1%) | 185 (23.1%) |
| **Subcutis Infiltration** | 0 (0.0%) | 20 (2.5%) | 0 (0.0%) | 780 (97.5%) | 20 (2.5%) | 0 (0.0%) |
| **PRAME Status: 0** | 54 (6.8%) | 174 (21.8%) | 77 (9.6%) | 495 (61.9%) | 228 (28.5%) | 131 (16.4%) |
| **PRAME Status: 1** | 9 (1.1%) | 42 (5.2%) | 69 (8.6%) | 680 (85.0%) | 51 (6.4%) | 78 (9.8%) |
| **PRAME Status: 2** | 13 (1.6%) | 26 (3.2%) | 186 (23.2%) | 575 (71.9%) | 39 (4.9%) | 199 (24.9%) |
| **PRAME Status: 3** | 9 (1.1%) | 38 (4.8%) | 117 (14.6%) | 636 (79.5%) | 47 (5.9%) | 126 (15.8%) |
| **PRAME Status: 4** | 183 (22.9%) | 252 (31.5%) | 83 (10.4%) | 282 (35.2%) | 435 (54.4%) | 266 (33.2%) |
| **Ki-67 Index: Low** | 100 (37.3%) | 64 (23.9%) | 22 (8.2%) | 82 (30.6%) | 164 (61.2%) | 122 (45.5%) |
| **Ki-67 Index: Intermediate** | 40 (14.9%) | 36 (13.4%) | 56 (20.9%) | 136 (50.7%) | 76 (28.4%) | 96 (35.8%) |
| **Ki-67 Index: High** | 20 (7.5%) | 8 (3.0%) | 30 (11.2%) | 210 (78.4%) | 28 (10.4%) | 50 (18.7%) |

Note: Confusion matrix of human-AI agreement. TP (True Positive) and TN (True Negative) indicate consensus; FN (False Negative) represents features identified by humans only; FP (False Positive) represents features identified by AI only. Marginal totals (Total Human GT / Total AI Pred) provide the overall frequency of feature detection. AI classifications for morphological criteria were optimized via probability-calibrated logistic regression models. Percentages for Ki-67 reflect only evaluable cases.

Supp. Table S4: Comparative Odds Ratios for Malignancy Prediction: Human vs. AI.

| **Criterion** | **Human GT: OR [95% CI]** | **Human GT: p** | **AI Pred: OR [95% CI]** | **AI Pred: p** |
|---|---|---|---|---|
| Dermal Mitoses | 1.23e+08 [1.31e-11 - 7.21e+222] | 0.989 | 3.83 [0.63 - 24.88] | 0.151 |
| Ulceration | 4.02e+06 [1.05e-87 - NA] | 0.993 | 1.57 [0.40 - 6.14] | 0.517 |
| Upward Melanocytes | 15.35 [7.14 - 35.74] | **< .001** | 1.80 [0.60 - 5.38] | 0.292 |
| Junctional Atypia | 12.42 [6.40 - 25.10] | **< .001** | 16.54 [4.60 - 62.58] | **< .001** |
| Pagetoid Spread | 4.34 [1.86 - 10.94] | **0.001** | 7.37 [1.78 - 31.79] | **0.006** |
| Dermal Atypia | 2.96 [0.86 - 10.01] | 0.081 | 7.26 [1.57 - 35.00] | **0.012** |
| Junctional Mitoses | 2.33 [0.42 - 43.63] | 0.430 | 0.89 [0.23 - 3.46] | 0.872 |
| Regression | 2.18 [0.87 - 5.84] | 0.108 | 0.81 [0.23 - 2.82] | 0.748 |
| Lack of Maturation | 1.36 [0.36 - 5.30] | 0.657 | 0.32 [0.06 - 1.79] | 0.196 |
| Atypical Dissociation | 0.47 [0.18 - 1.19] | 0.113 | 0.07 [0.01 - 0.48] | **0.007** |
| Nest Confluence | 0.44 [0.22 - 0.87] | **0.020** | 0.78 [0.30 - 2.03] | 0.618 |
| Symmetry | 0.34 [0.17 - 0.66] | **0.002** | 0.15 [0.03 - 0.61] | **0.009** |
| Ordered Single Cells | 0.26 [0.13 - 0.51] | **< .001** | 0.19 [0.09 - 0.41] | **< .001** |

Note: Multivariate logistic regression comparing the predictive impact of individual features on malignancy. 'Human GT' refers to the model based on pathologist-defined ground truth; 'AI Pred' refers to the model based on AI-derived feature probabilities. Bold p-values indicate statistical significance ($p < 0.05$).

### Supplementary Material S2

**Standardized Histopathological Annotation Catalog for Melanocytic Lesions (Version 4.0)**

**Introduction:** This catalog defines the histological criteria assessed for each case in this study. All evaluations were performed by a board-certified dermatologist and dermatopathologist. The assessment integrated Hematoxylin and Eosin (H&E) slides and any available immunohistochemical (IHC) stains (including cases where PRAME, SOX10, Melan-A, HMB45, or Ki-67 had been performed). The objective of this standardized catalog is to ensure consistent annotation for both statistical modeling and computational pathology applications.

---

#### I. Structural Top-Level Criteria

*(These criteria determine the applicability of subsequent sections)*

**1.0 Junctional Component Present? Yes / No** *Definition:* The lesion shows a melanocytic proliferation at or along the dermo-epidermal junction. Purely dermal lesions are scored as "No."

| | |
|---|---|
| 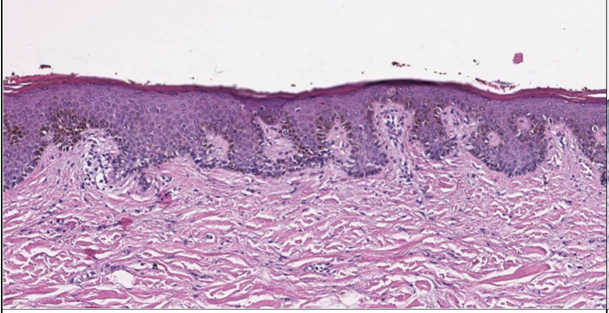 | 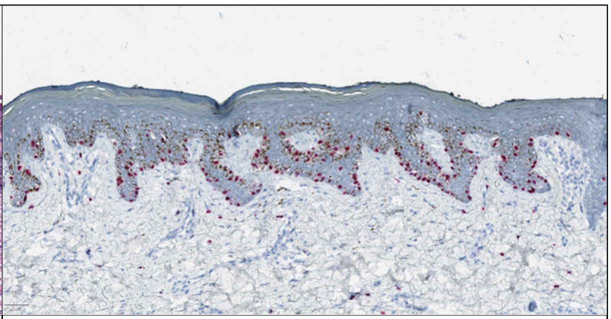 |
| Yes (H&E) | Yes (Sox10) |

**2.0 Dermal Component Present? Yes / No** *Definition:* The lesion contains melanocytes within the dermis that are not associated with adnexal structures (e.g., hair follicles). Purely junctional lesions (e.g., melanoma in situ) are scored as "No."

| | |
|---|---|
| 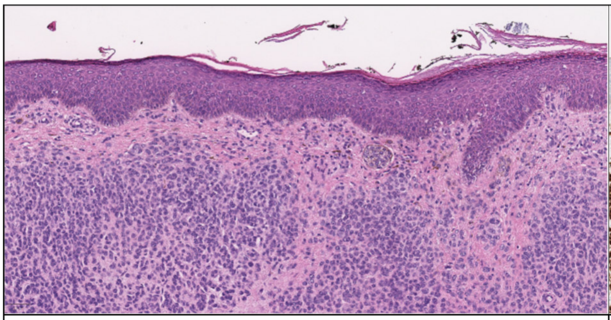 | 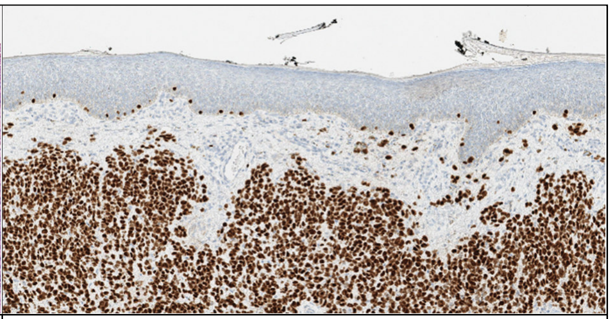 |
| Yes (H&E) | Yes (Sox10) |

---

#### A. Low-Power Architecture (H&E)

**A.1 Symmetry: Yes / No** *Definition:* **"Yes"** (i.e., symmetrical) is annotated if the lesion, viewed at low magnification, exhibits a balanced and approximately mirror-image distribution of mass, shape, and pigmentation across an imaginary vertical midline. **"No"** (i.e., asymmetrical) is annotated if the two halves of the lesion show significant disparities in these parameters.

| 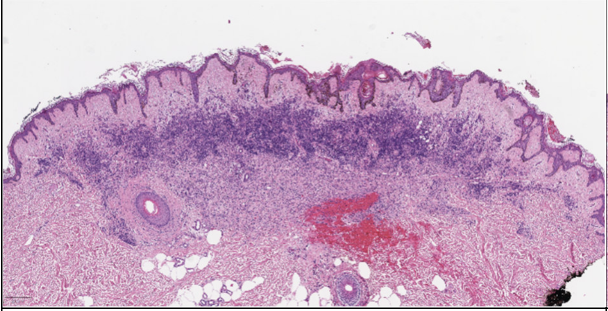 | 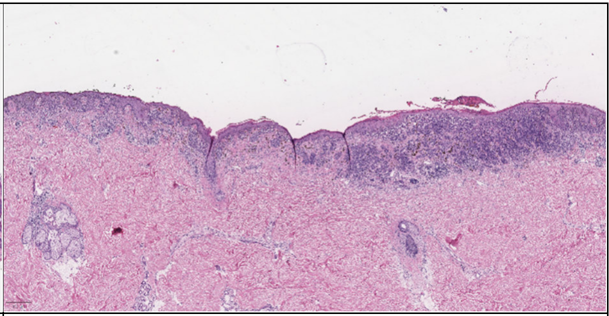 |
| --- | --- |
| Yes (H&E) | No (H&E) |

#### B. Epidermis & Junctional Zone

*(Assessed using H&E, supplemented by available melanocytic markers such as SOX10 for enhanced visualization when performed)*

**B.1 Upward Melanocytic Migration: Yes / No** *Definition:* "Yes" is annotated if melanocytes are identified above the basal layer (suprabasal, within the stratum spinosum or higher). This is an inclusive criterion that ranges from isolated, scattered single cells (often highlighted by IHC) to the extensive involvement described in B.2. Every case meeting the criteria for Pagetoid Spread (B.2) is by definition also positive for B.1.

| 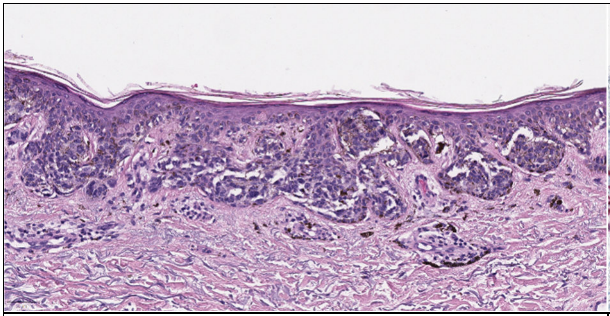 | 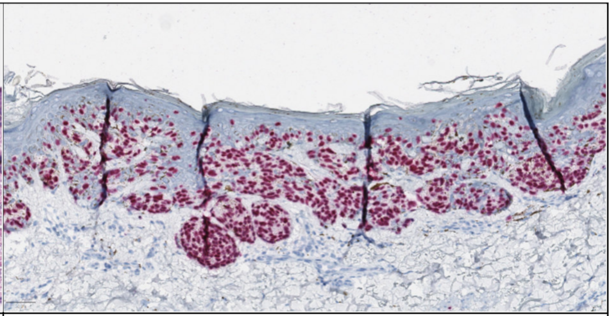 |
| --- | --- |
| Yes (H&E) | Yes (Sox10) |

**B.2 Pagetoid Spread: Yes / No** *Definition:* "Yes" is annotated if there is a conspicuous, high-density, or nest-like distribution of melanocytes within the upper two-thirds of the epidermis. This represents a qualitative and quantitative escalation of upward migration, characterized by disseminated involvement or the presence of intraepidermal melanocytic nests.

| 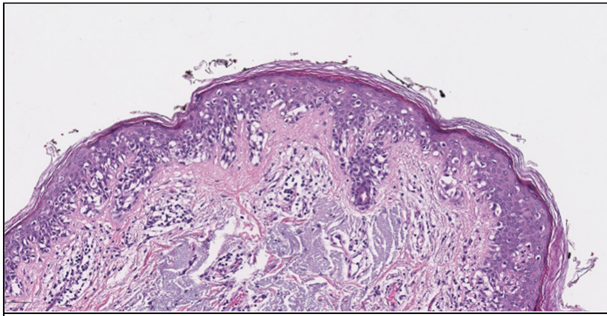 | 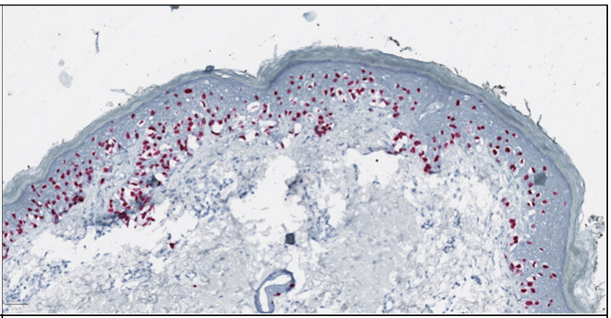 |
| --- | --- |
| Yes (H&E) | Yes (Sox10) |

**B.3 Confluence of Junctional Nests: Yes / No** *Definition:* "Yes" is annotated if adjacent junctional melanocytic nests merge horizontally, leading to "bridging" of the rete ridges.

| | |
|---|---|
| 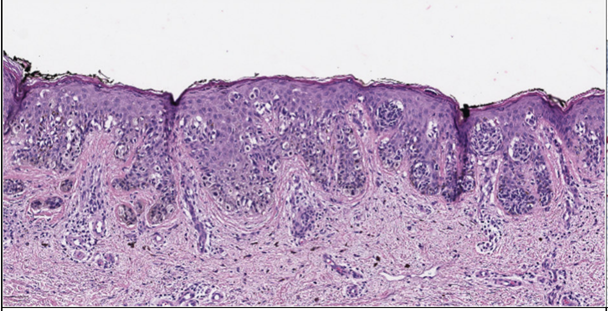 | 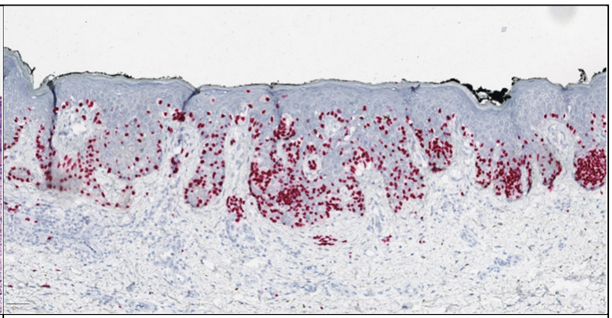 |
| Yes (H&E) | Yes (Sox10) |

**B.4 Marked Junctional Atypia: Yes / No** *Definition:* "Yes" is annotated if junctional melanocytes exhibit features of cytological atypia. This assessment is based on a combination of the following:

- **Nuclear enlargement:** Nuclei significantly larger than those of adjacent basal keratinocytes (macrokaryosis).
- **Nuclear contour irregularities:** Jagged, indented, or non-oval nuclear membranes (notching).
- **Pleomorphism:** Pronounced variation in nuclear size and shape.
- **Chromatin changes:** Hyperchromasia or coarse, clumpy chromatin distribution.
- **Nucleolar prominence:** Presence of enlarged or conspicuous nucleoli.

| |
|---|
| 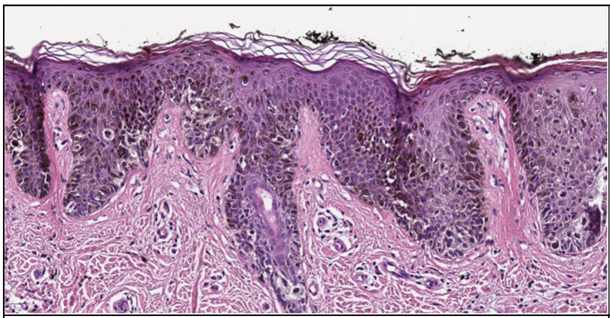 |
| Yes (H&E) |

**B.5 Atypical Junctional Mitoses: Yes / No** *Definition:* "Yes" is annotated if at least one clearly non-canonical mitotic figure (e.g., tripolar, multipolar, or asymmetrical) is identified within the junctional component.

| | |
|---|---|
| 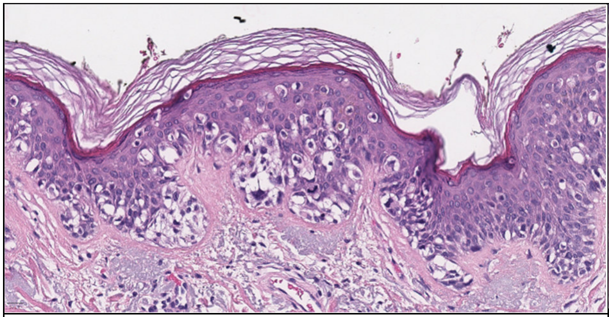 | 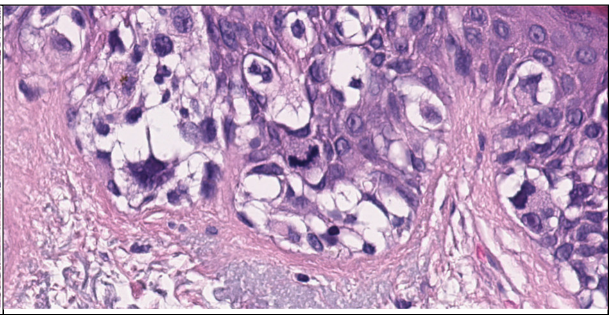 |
| Yes (H&E) | Zoomed in left picture |

**C. Dermis**

*(Assessed using H&E, supplemented by available melanocytic markers such as SOX10 for enhanced visualization when performed)*

**C.1 Lack of Maturation (within nests): Yes / No** *Definition:* "Yes" is annotated if melanocytes organized in nests or clusters in the deep dermal component show no reduction in cellular or nuclear size.

| 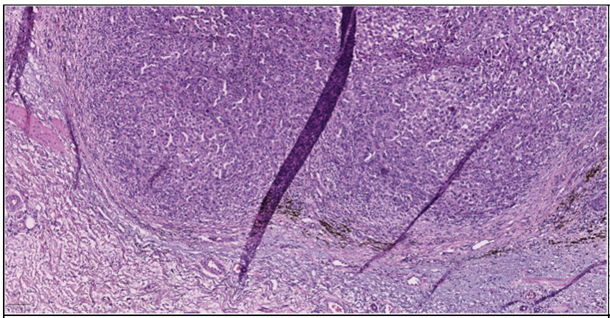 | 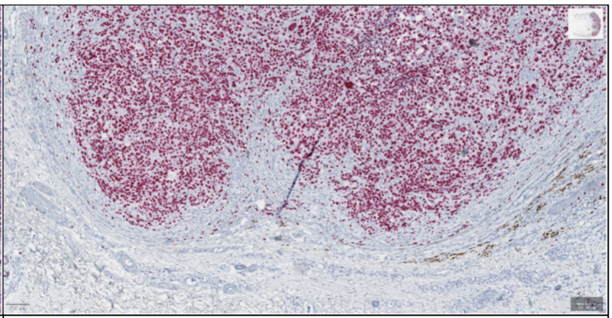 |
|---|---|
| YES (H&E) | YES (Sox10) |

**C.2 Marked Dermal Atypia: Yes / No** *Definition:* "Yes" is annotated if dermal melanocytes meet the criteria for marked atypia as described in section B.4.

| 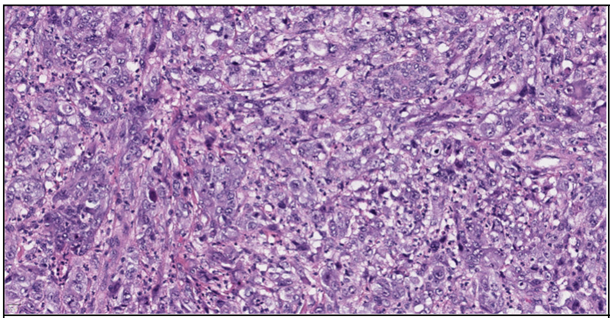 |
|---|
| YES (H&E) |

**C.3 Dermal Mitoses: Yes / No** *Definition:* "Yes" is annotated if at least one distinct mitotic figure (typical or atypical) is identified within the dermal tumor component.

| 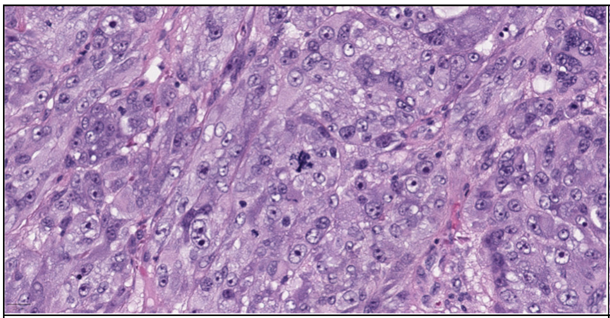 | 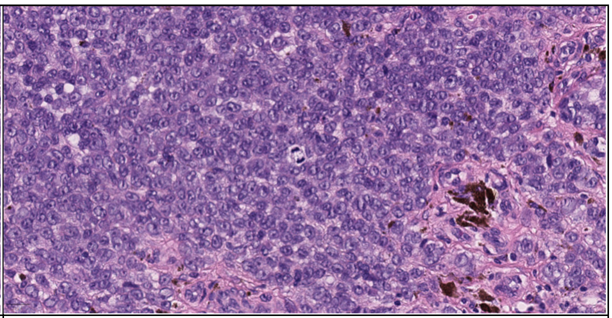 |
|---|---|
| Yes (H&E) | Yes (H&E) |

**C.4 Ordered Single-Cell Pattern (Benign Maturation): Yes / No** *Definition:* "Yes" is annotated if the deep dermis shows an orderly arrangement of individual, small, uniform (spindled or nevus-like) melanocytes interspersed between collagen bundles. This pattern is a classical sign of physiological maturation (Type-C cells).

| |
|---|
| 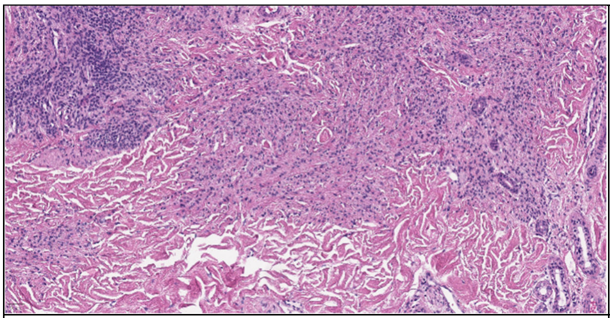 |
| Yes (H&E) |

**C.5 Atypical Single-Cell Dissociation (Malignant Pattern): Yes / No** *Definition:* "Yes" is annotated if individual, cytologically atypical tumor cells are found within the dermal stroma without nest formation, suggesting a pathological loss of cell-cell adhesion.

| | |
|---|---|
| 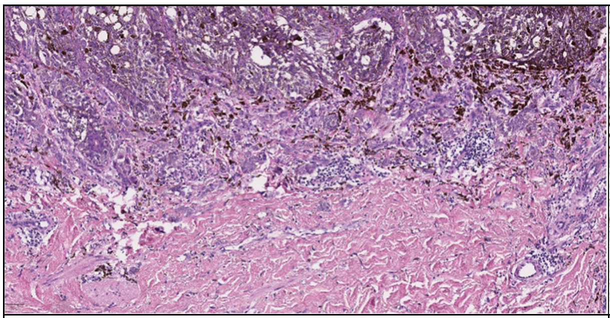 | 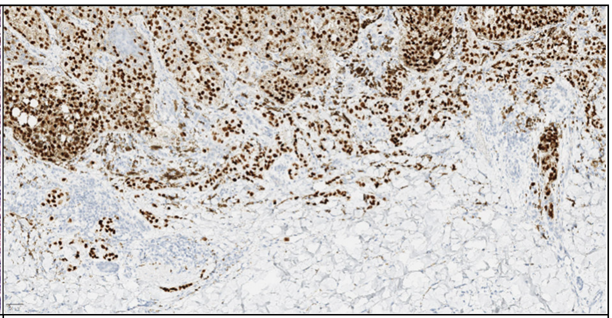 |
| Yes (H&E) | Yes (Sox10) |

**C.6 Dense Lymphocytic Infiltrate: Yes / No** *Definition:* "Yes" is annotated if a dense, band-like, or diffuse lymphocytic infiltrate is present, accompanying or infiltrating the lesion and largely obscuring the normal collagen.

| | |
|---|---|
| 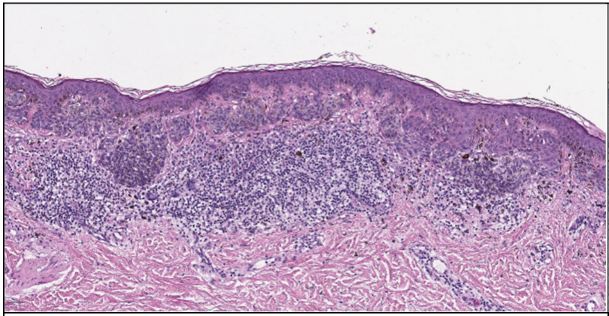 | 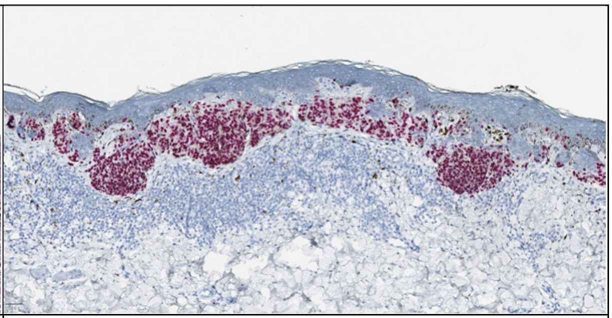 |
| Yes (H&E) | Yes (Sox10) |

**C.7 Intratumoral Lymphocytes (TILs): Yes / No** *Definition:* "Yes" is annotated if lymphocytes are identified in direct contact with tumor cells within the dermal nests or tumor clusters.

| | |
|---|---|
| 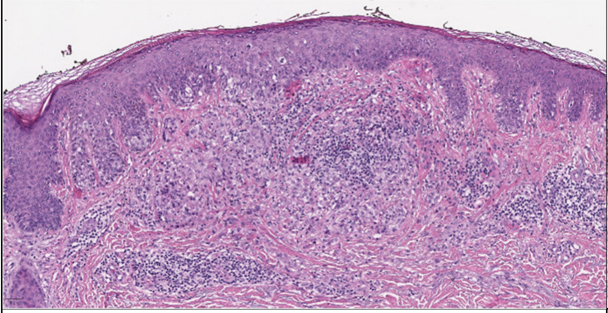 | 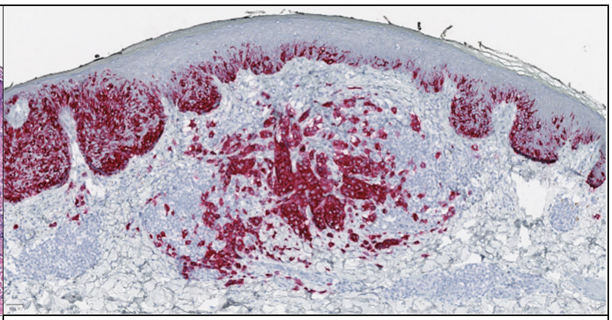 |
| Yes (H&E) | Yes (Sox10) |

**C.8 Ulceration: Yes / No** *Definition:* "Yes" is annotated if there is a focal or broad-based loss of the epidermis with underlying fibrin and neutrophil infiltration in direct association with the lesion.

| |
|---|
| 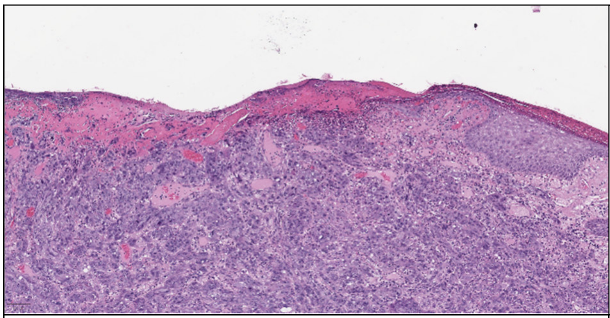 |
| Yes (H&E) |

**C.9 Regression-like Fibrosis: Yes / No** *Definition:* "Yes" is annotated if a focal or diffuse area of the superficial dermis exhibits horizontal fibrosis (cicatrization), often accompanied by increased small-caliber vascularity (neocapillarization) and/or the presence of pigment-laden macrophages (melanophages). For the purpose of this blinded evaluation, this category encompasses all areas of dermal scarring, as a definitive distinction between spontaneous biological regression and post-interventional scarring cannot be reliably made without clinical history.

| |
|---|
| 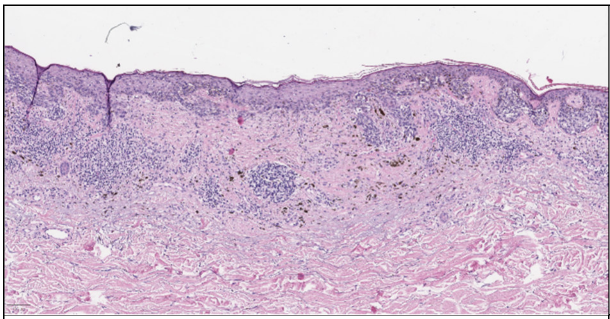 |
| Yes (H&E) |

##### D. Subcutis

**D.1 Subcutaneous Infiltration: Yes / No** *Definition:* "Yes" is annotated if tumor cells clearly infiltrate the subcutaneous adipose tissue.

| |
|---|
| 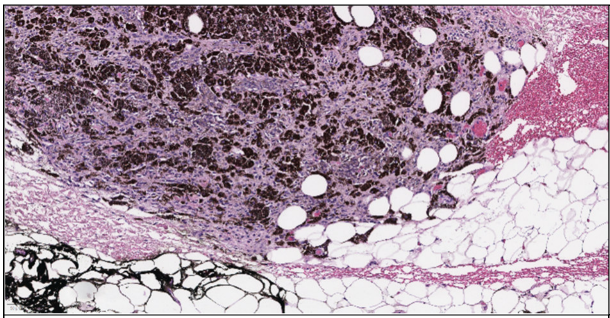 |
| Yes (H&E) |

---

##### E. Specific Immunohistochemical Markers

**E.1 Ki-67 (Mib1) Proliferation Index: Low (<5%) / Intermediate (5–20%) / High (>20%)** *Definition:* Visual estimation of the percentage of positively stained tumor cell nuclei in the region of highest proliferative activity ("hotspot").

| | |
|---|---|
| 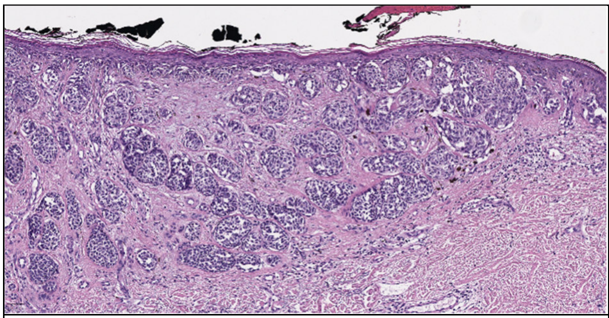 | 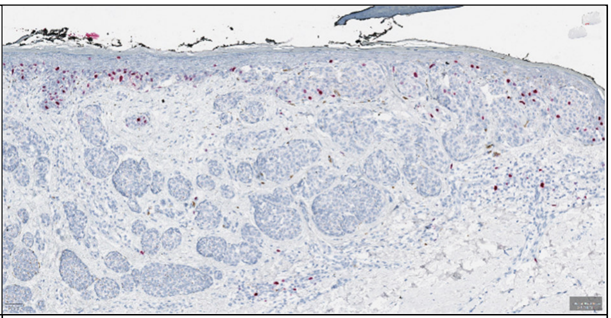 |
| Ki-67 (Mib1): Low (<5%); H&E | Ki-67 (Mib1): Low (<5%); IHC |
| 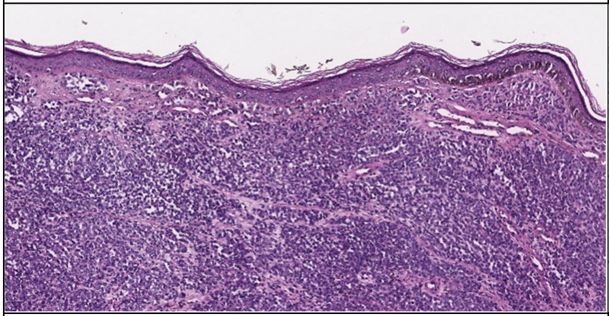 | 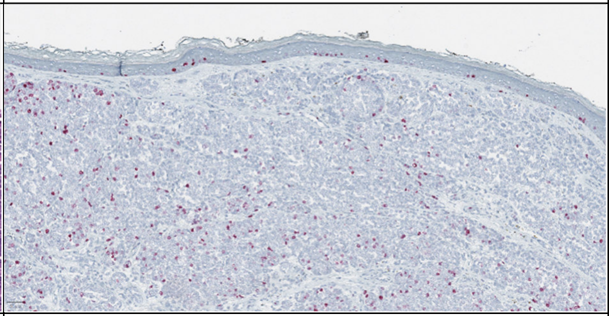 |
| Ki-67 (Mib1): Intermediate (5–20%); H&E | Ki-67 (Mib1): Intermediate (5–20%); IHC |
| 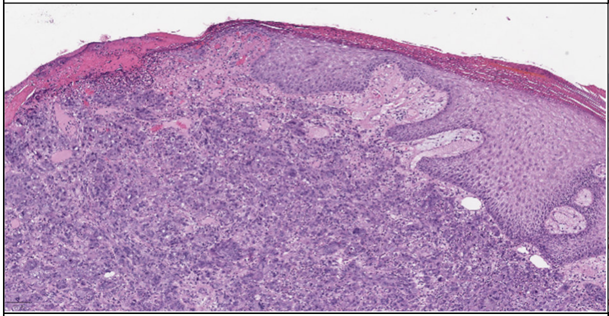 | 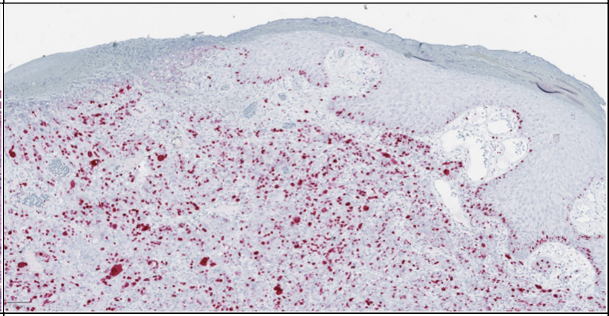 |
| Ki-67 (Mib1): High (>20%); H&E | Ki-67 (Mib1): High (>20%); IHC |

**E.2 PRAME Status: 0 / 1+ / 2+ / 3+ / 4+** *Definition:* Assessment of nuclear PRAME expression across the entire melanocytic tumor tissue according to the following scale:

- **0:** No positive nuclei
- **1+:** 1–25% of tumor cell nuclei are positive
- **2+:** 26–50% of tumor cell nuclei are positive
- **3+:** 51–75% of tumor cell nuclei are positive
- **4+:** >75% of tumor cell nuclei are positive

| | |
|---|---|
| 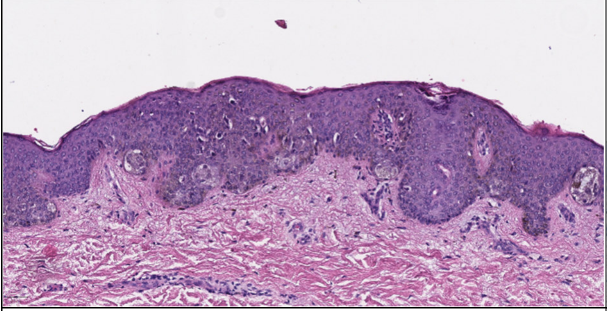 | 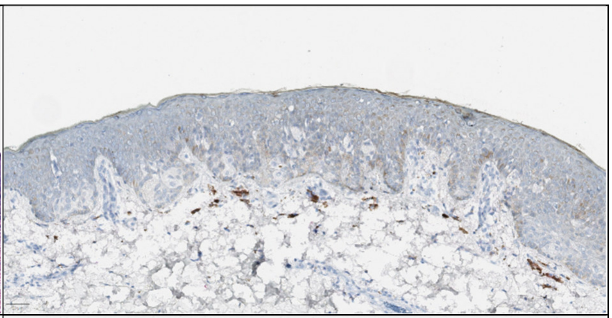 |
| **Prame 0**: No positive cells<br>H&E upper left<br>Prame upper right<br>Sox10 lower right | 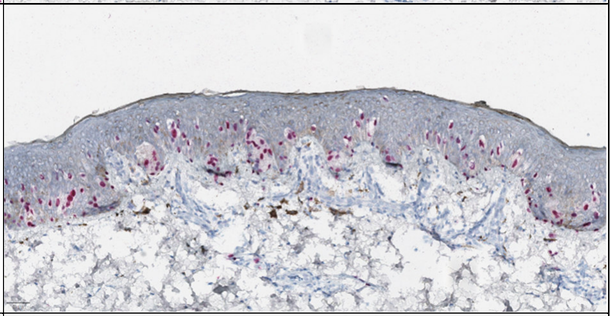 |
| 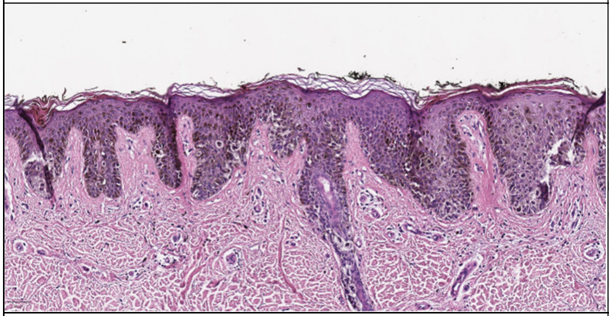 | 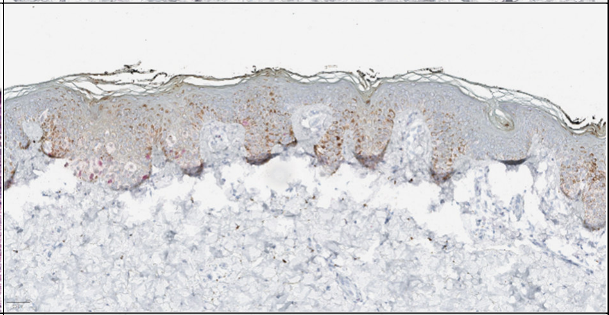 |
| **Prame 1+**: 1–25% positive cells<br>H&E upper left<br>Prame upper right<br>Sox10 lower right | 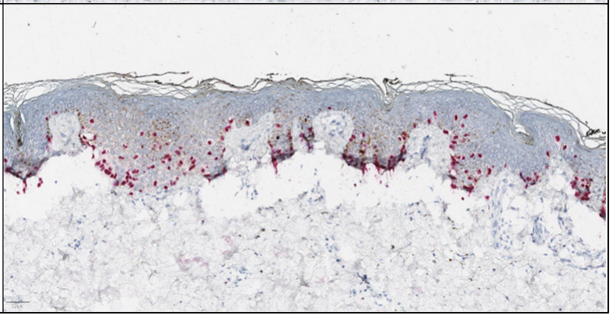 |

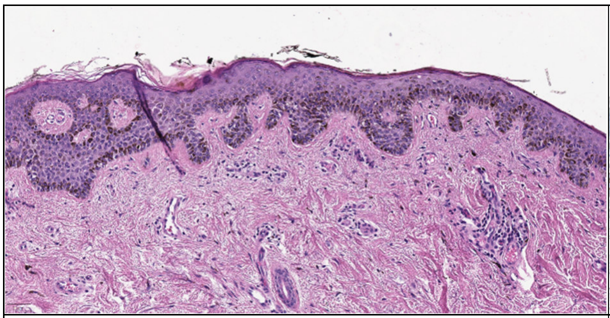

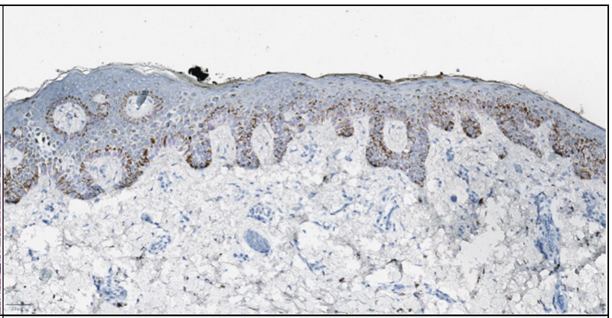

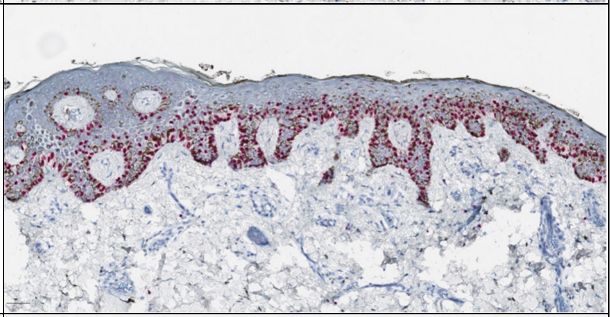

**Prame 2+**: 26–50% positive cells
H&E upper left
Prame upper right
Sox10 lower right

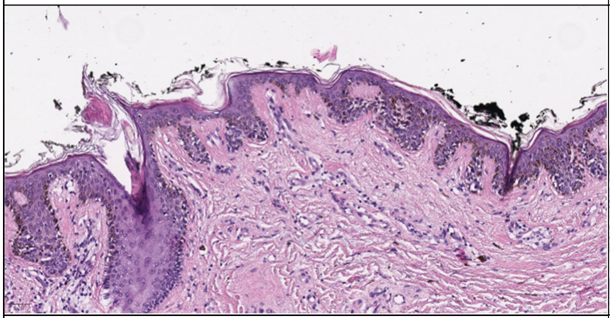

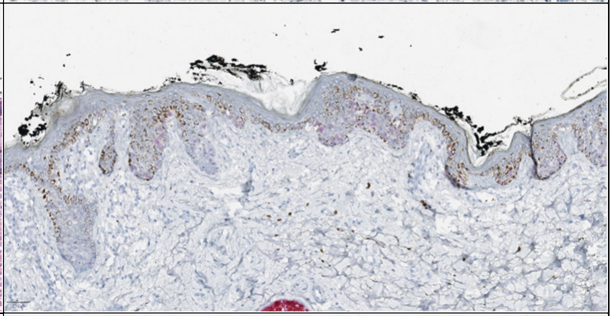

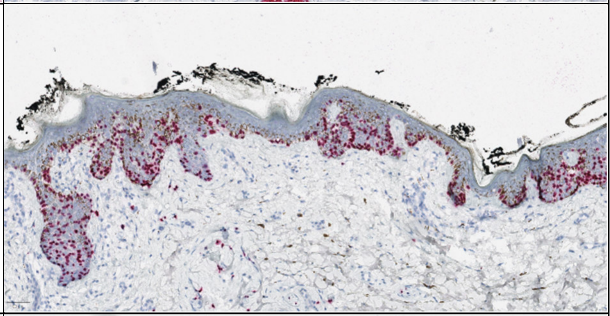

**Prame 3+**: 51–75% positive cells
H&E upper left
Prame upper right
Sox10 lower right

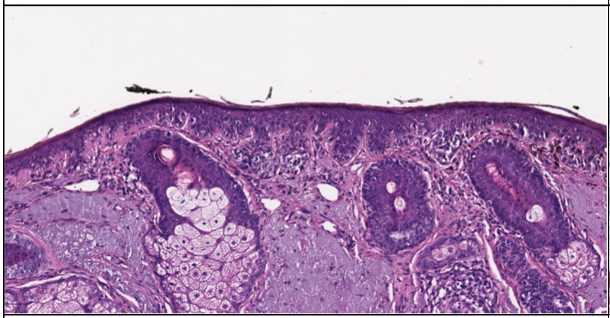

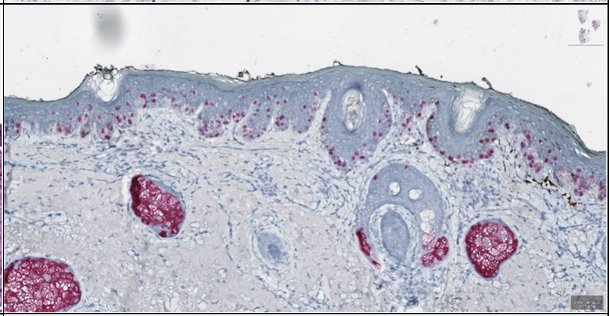

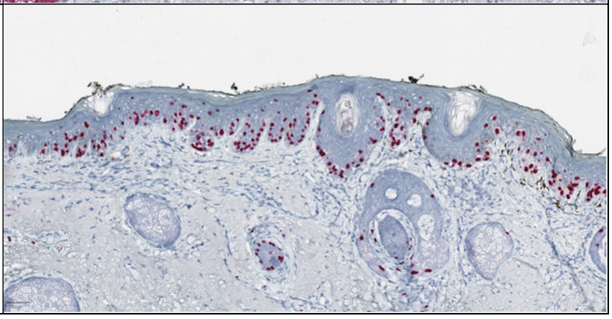

**Prame 4+**: >75% positive cells
H&E upper left
Prame upper right
Sox10 lower right
